# Profiling functional networks identify activation of corticostriatal connectivity in ET patient after MRgFUS thalamotomy

**DOI:** 10.1101/2023.11.22.23298885

**Authors:** Jiaji Lin, Xiaopeng Kang, Jiayou Zhou, Dekang Zhang, Jianxing Hu, Haoxuan Lu, Longsheng Pan, Xin Lou

**Author notes:** These authors contributed equally to this work (Jiaji Lin, Xiaopeng Kang, Jiayou Zhou). Correspondence to: Xin Lou, Department of Radiology, Chinese PLA General Hospital/Medical School of Chinese PLA, No.28 Fuxing Road, Beijing, 100853, China., Longsheng Pan, Department of Neurosurgery, Chinese PLA General Hospital/Medical School of Chinese PLA, No.28 Fuxing Road, Beijing, 100853, China.

## Abstract

**Objective:** MR-guided focused ultrasound (MRgFUS) thalamotomy is a novel and effective treatment for medication-refractory tremor in essential tremor (ET), but how the brain responds to this deliberate lesion is not clear. The current study aimed to evaluate the immediate and longitudinal alterations of functional networks after MRgFUS thalamotomy.

**Methods:** We retrospectively obtained preoperative and postoperative 30-day, 90-day, and 180-day data of 31 ET patients subjected with MRgFUS thalamotomy from 2018 to 2020. Their archived resting-state functional MRI data were used to functional network comparison as well as graph-theory metrics analysis. Both partial least squares (PLS) regression and linear regression were conducted to associate functional features to tremor symptoms.

**Results:** MRgFUS thalamotomy dramatically abolished tremors, while global functional network only sustained immediate fluctuation within one week postoperatively. Network-based statistics have identified a long-term enhanced corticostriatal subnetwork by comparison between 180-day and preoperative data (*P* = 0.019). Within this subnetwork, network degree, global efficiency and transitivity were significantly recovered in ET patients right after MRgFUS thalamotomy compared to the pre-operative timepoint (*P* < 0.05), as well as hemisphere lateralization (*P* < 0.001). The PLS main component significantly accounted for 33.68% and 34.16% of the total variances of hand tremor score and clinical rating scale for tremor (CRST)-total score (*P* = 0.037 and 0.027). Network transitivity of this subnetwork could serve as a reliable biomarker for hand tremor score control prediction at 180-day postoperatively (β = 2.94, *P* = 0.03).

**Conclusions:** MRgFUS thalamotomy promoted corticostriatal connectivity activation correlated with tremor improvement in ET patient after MRgFUS thalamotomy.

## 1. Introduction

Magnetic resonance-guided focused ultrasound (MRgFUS) is a novel and promising technology that allows the delivery of spherical-phased converging beams to a specific brain target using MR imaging (MRI) for guidance [1]. Thanks to its advantages such as no need for craniotomy, anesthesia, or ionizing radiation, MRgFUS application has led to a renaissance of brain lesion therapy, such as thalamotomy for tremor disease [2]. Multiple studies have presented that MRgFUS thalamotomy can precisely ablate ventral intermedius nucleus (Vim) and sustained > 78% tremor relieve for intractable essential tremor (ET) and >85% tremors improvement for tremor-dominant Parkinson’s disease (PD)[3]. Thus far, the safety of MRgFUS thalamotomy has been widely proven and become increasingly for the patients with various types of tremor in the United States, Japan, South Korea, and China.

With its potential wide clinical application, there is an increasing interest regarding the immediate and longitudinal effect of MRgFUS thalamotomy on brain functional activity. Jang et al. examined pre-supposed functional motor-tremor networks alteration after in ET patients, and found that MRgFUS thalamotomy regulates interactions over the motor network via symptom-related connectivity changes but accompanied by transient, symptom-unrelated diaschisis in the brain network [4]. Our recent study has indicated a significant perturbation in the macroscale gradient frameworks post MRgFUS thalamotomy with rebalance of functional hierarchical architecture [5]. These results suggest that brain integrity violation by Vim ablation may promote a continuous reconstruction of functional steady state, involving mechanisms such as lesion absorption, neurotransmitter regulation and so on. In fact, clues have been found in much earlier studies of structural wirings: diffusion-tensor MRI study of Zur et al. investigated the effects of MRgFUS thalamotomy on fiber tract integrity, finding long-term damage in the ablated core and from the thalamus to the red nucleus tract [6]. Diffusion imaging also suggested continuous connectivity alteration related to subthalamic nucleus, associated both with clinical outcomes of MRgFUS thalamotomy [7, 8]. Network-based statistics identified a U-shaped alteration in regional tract communication after MRgFUS thalamotomy, accompanied by relevant cerebral blood flow and gray matter adaptation [9]. All these pose new challenges to us: a more comprehensive and macroscopic understanding of the postoperative alteration spectrum are of great significance for us to further identity an ideal time window for functional research for better understand and prediction of the benefits and side effects of MRgFUS thalamotomy.

To this end, we collected longitudinal resting-state functional MRI scanning up to half year post MRgFUS thalamotomy in ET patients to investigate their altered functional features and the correlation with tremor symptoms.

## 2. Methods

### 2.1 Study design and registration

The present study is retrospective, which was registered in clinicaltrials.gov with NCT04570046 to investigate the functional MR images related to MRgFUS thalamotomy in Chinese PLA General Hospital from 2018 to September 2020. It was approved by the institutional review board and the independent scientific advisory committee at Chinese PLA General Hospital. The data were anonymous, and the requirement for informed consent was therefore waived. Briefly, the image data of ET patients were enrolled in clinical trial of NCT03253991, which was an interventional study to evaluate the effectiveness and safety of ExAblate MRgFUS in 2017. Until September 2020, a total of 31 of these patients successfully completed preoperative and postoperative of imaging follow-up and tremor assessment, and all these data were collected for the retrospective study. Resting state functional MRI scanning was performed pre-operatively, and at 1-day, 7-day, 30-day, 90-day and 180-day postoperatively. Additional data of 31 healthy controls with matched the sex, age and education with ET patients were collected during the same period.

### 2.2 MRgFUS thalamotomy and tremor assessment

All subjects were right-handed according to the Edinburgh Handedness Scale [10] and have unilateral tremor manifestations on the right extremity. Unilateral MRgFUS thalamotomy in left Vim was performed in a 3T MRI suite (Discovery 750, GE Healthcare, USA) using the ExAblate Neuro focused ultrasound system as previous reports (InSightec, Israel) [9]. Briefly, the patient’s head was fixed, and low-power sonication was used for the initial target ablation and adjusted based on real-time patient feedback. After final target was determined, the energy was increased so that the target tissue ablation temperature reached approximately 55-60°C.

Tremor performance was assessed in off-medication state using Clinical Rating Scale for Tremor (CRST) ratings preoperatively and at 30-day, 90-day, and 180-day after MRgFUS thalamotomy, including CRST-A assesses tremor location and amplitude, CRST-B assesses tremor when performing specific motor tasks, and CRST-C assesses functional disability due to tremor [9]. Primary hand tremor score contralateral to the left Vim was measured using a derived subscale composed of CRST parts A (three items on a scale of 0-4 for each: rest, posture, and action components of hand tremor) and B (five tasks on a scale of 0-4 for each: handwriting-dominant hand only, drawings and pouring). The hand tremor improvement ratio was calculated with the formula: (postoperative scores - preoperative scores) / preoperative scores × 100%.

### 2.3 MRI acquisition and preprocessing

MRI data were collected on a Discovery MR750 scanner (General Electric, Milwaukee, WI). Patients were instructed to relax, keep their eyes closed, not sleep or think specifically during MRI acquisition. Ear plugs and tight but comfortable foam padding were used to reduce noise and head motion. The resting state functional MRI data were acquired using an echo-planar imaging (EPI) sequences with the following parameters: repetition time (TR) = 2000 ms; echo time (TE) = 30 ms; flip angle (FA) = 90°; field of view (FOV) = 240 mm × 240 mm; matrix = 64 × 64; slice thickness = 3.5 mm; slice gap = 0.5mm; interleaved slices = 36. Each functional run produced 180 volumes. The high-resolution structural data were obtained using a three-dimensional fast spoiled gradient recalled (3D FSPGR) sequence with the following parameters: TR = 6.656 ms; TE = 2.928 ms; inversion time = 800 ms; FA = 7°; FOV = 265 mm × 265 mm; matrix = 256 × 256; slice thickness = 1 mm; and contiguous sagittal slices = 192.

The preprocessing procedures of all EPI image were implemented using GRETNA toolkits [11]. Briefly, the first five time points were removed for signal equilibrium and participants’ adaptation. Slice timing, head motion correction, and spatial normalization to standard space were performed. A sample-specific diffeomorphic image registration through the exponentiated Lie algebra (DARTEL) template was created using structural images [12]. The EPI volumes were then normalized, resampled and smoothed with kernel of 2 times the voxel size. Covariates were regressed out from the time series of every voxel. The covariates included a total of 26 variables, including the white matter signal, the cerebrospinal fluid signal, and the Friston 24-parameter model.

### 2.4 Function network construction and subnetwork extraction

Linear detrending and a temporal filter (0.01-0.1 Hz) were applied to reduce low-frequency drifts and high frequency physiological noise. As our recent study [5], all functional connectomes were constructed by an intrinsic functional connectivity atlas (Table S1 or https://github.com/louxin-lab). Pearson correlation coefficients were computed for each pair of brain regions in the intrinsic functional connectivity atlas [13], and then combined into 1,039 × 1,039 undirected and weighted connectivity matrices as a global functional network. Considering damage to the hemisphere integrity by MRgFUS thalamotomy, hemispheres functional activity was analyzed [14, 15]. All raw connectivity values were summed up according to the hemisphere distribution (right / left hemisphere functional activity = both brain regions pair of the functional connectivity are in the right / left hemisphere), and hemisphere lateralization was calculated with the formula: hemisphere lateralization = right hemisphere functional activity / left hemisphere functional activity. Moreover, a Fisher r-to-z transform was computed to normalize raw connectivity values for group comparison. Connectivity alterations were measured by two-tail *t* test between the ET patients and matched health controls (HCs), and all alterations collected for distribution analysis.

A non-parametric connectome-wide method called network-based statistic was applied to identify disconnected regional subnetworks[16]. Fisher z-transformed global function networks were applied and over 5000 permutations were performed for global functional network between Pre-op and postoperative 180-day. A *P* < 0.05 threshold was applied. Both pyecharts (https://github.com/pyecharts/pyecharts) and circos [17] were applied to visualize the subnetwork analysis. Furthermore, rich-club organization was characterized by a high degree of above-average interconnected nodes, beyond that expected by chance and provides a basic substrate for integrating and disseminating information across the whole network [18, 19]. The edges in the regional subnetwork were binarized, and the degree of each node was calculated which was defined as the number of connections of this node. The top 5 nodes with the highest degree were taken as the rich clubs of the regional subnetwork.

### 2.5 Graph-theory metrics analysis

Next, we explored the topological properties in functional networks. The raw connectivity matrices were absolutized and top 10% thresholded, and then underwent graph-theory network analyses using the Brant toolbox [20]. The network degree was applied for the overall and basic measure of the functional wiring and coherence. Global efficiencies were calculated separately to analyze network functional integration, which characterizes the efficiency of parallel information transfer within a functional network or brain region [21]. Transitivity was used as measures of functional segregation. Transitivity is a classical variant of the clustering coefficient suitable for network analysis, which is collectively normalized and not disproportionately influenced by the network degree distribution[22]. In order to further analyze the alteration in anatomical regions (cortex, cerebellum, thalamus and striatum), the raw and weighted connectivity matrices were absolutized and all linked edges of the specific node were summed up as its degree. The regional degree is defined as the sum of all nodal degrees within this region [23, 24].

### 2.6 Spontaneous neuronal activity analysis

Amplitude of low-frequency fluctuation (ALFF), measures of spontaneous blood oxygen level dependent (BOLD) signal oscillations, have been proposed as a suggestive of spontaneous neuronal activity [25]. As a normalized index of ALFF, fractional ALFF (fALFF) can provide more sensitive and specific measures of spontaneous neuronal activity by suppressing non-specific signal components [26]. In present study, ALFF and fALFF were measured using RESTplus v1.24 software [27]. Briefly, after preprocessing, a temporary bandpass range as 0.01 - 0.08 Hz was set to filter the BOLD time series for each voxel to remove the very-low frequency and high frequency noise. Then the filtered time series were transformed to the frequency domain using a fast Fourier transformation to obtain the power spectrum. Next, ALFF for each voxel was calculated as the averaged square-rooted power spectrum within the frequency band of 0.01 - 0.08 Hz. Finally, a division of ALFF within the specified frequency band by the entire frequency range was computed at each voxel to yield the fALFF. Both ALFF and fALFF maps were standardized into z-score maps (zALFF and zfALFF) to minimize the global effects of variability.

### 2.7 Symptom prediction by functional features

In order to analyze the correlation between the functional features of MRgFUS thalamotomy subnetwork and the primary tremor symptoms, we introduced Partial least squares (PLS) regression to investigate their optimal predictive relationship as in Whitaker and Seidlitz [28, 29]. This multivariate technique seeks to find the latent variables (PLS components) which maximize the correlation between predictor variables and response variables. Here, multiple tremor ratings were used as response variables, and raw connectivity values of subnetwork in ET patients were used as predictor variables. Intrinsic permutations tests were performed at *P* < 0.05 significance threshold for general variance explanation of the principle PLS components. The PLS weights based on functional connectivities contribution to PLS components were applied to tremor symptom prediction. Additional Pearson correlation was applied to verify the agreement between predicted ratios and actual results. Moreover, graph-theory metrics and hemisphere lateralization features were also used to compared Pearson correlation coefficients with primary tremor symptoms all well. The features with a significance < 0.05 were considered significant.

We also test whether pre-operative features of functional features of MRgFUS thalamotomy subnetwork can provide a prediction for right tremor improvement ratio at 180-day postoperatively. Supervised pattern learning was applied to associate global gradient features to tremor symptom. All the raw functional connectivities of MRgFUS thalamotomy subnetwork were used to predict the tremor ratings by support vector regression. Training and performance evaluation employed nested three folds cross-validation with 10,000 repetitions. In the testing, the remaining test fold was used to measure prediction performance. We repeated the training-test procedure across all folds for median performance. Predicted tremor symptoms were compared using Pearson correlation coefficients and mean absolute error (MAE). A total of 10,000 permutation tests with randomly shuffled tremor ratings determined whether our prediction exceeded the chance level. Moreover, predictive performances of graph-theory metrics were also evaluated for tremor improvement ratios at postoperative 6-month by linear models. Network degree, global efficiencies and transitivity of MRgFUS thalamotomy subnetwork were included into a linear regression analysis with adjustment of subject’s sex, age, and constant.

### 2.8 Data and code availability

The conditions of our ethics approval do not permit public archiving of raw data. Quantile-quantile plots were used to evaluate the normality of the data. The linear mixed-effect model (LMM) was used to analyze the trend if necessary. Most computations were performed in the Python or R engine. All code and statistical image data are available at https://github.com/louxin-lab.

## 3. Results

There was a total of 31 medication-resistant ET patients who underwent unilateral MRgFUS thalamotomy were finally included in the present study. The mean age of these patients was 62.16 ± 10.88 years with disease duration of 18.03 ± 10.35 years. Among the patients, 21 patients were male, and 71.00% of patients had a family history of ET. Basic information of ET patients and HCs could be found in Table 1. At the preoperative baseline (Pre-op) of ET patients, there were significant tremor manifestations on the right extremity, with hand tremor scores of 21.23 ± 4.23 and CRST-total scores of 57.61 ± 11.54 (Table 2). MRgFUS thalamotomy resulted in significant relieve in tremor performance (LMM: *P*_for trend_ < 0.001 for both hand tremor score and CRST-total score)(Figure 1A). At 180-day post focused ultrasound treatment, these hand tremor scores decreased to 5.27 ± 5.86 points (Control ratio = 75.18%, *P* < 0.001 compared with Pre-op) and CRST-total scores decreased to 22.73 ± 16.78 (Control ratio = 60.55%, *P* < 0.001 compared with Pre-op). No patients experienced significant postoperative complications and side effects during the follow-up of 6 months.

**Figure 1.**
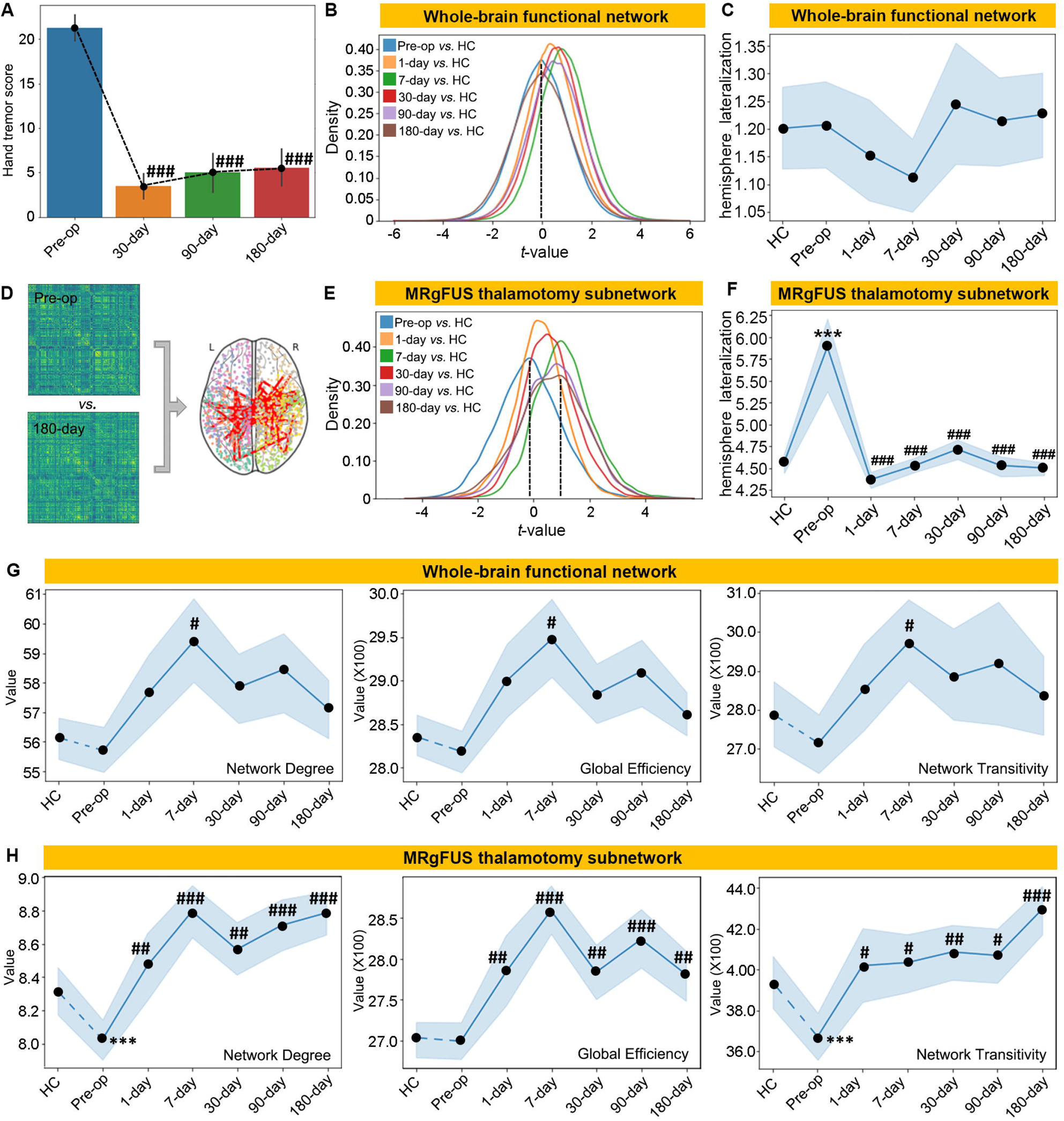
Global and regional brain networks presented different postoperative immediate and longitudinal alterations after MRgFUS thalamotomy. **(A)** Temporal changes of hand tremor score of ET patients after MRgFUS thalamotomy. **(B and E)** Distribution maps of functional connectivity alteration for the global functional network and MRgFUS thalamotomy subnetwork at different postoperative timepoints compared to matched HCs by *t*-test. **(C and F)** Temporal changes of hemisphere lateralization for the global functional network and MRgFUS thalamotomy subnetwork. **(D)** Network-based statistics were performed based on the global functional network between postoperative 180-day and Pre-op timepoint. **(G and H)** Temporal changes of network metrics for the global functional network and MRgFUS thalamotomy subnetwork. (Pre-op = pre-operative; 1-day = 1-day postoperatively; 7-day = 7-day postoperatively; 30-day = 30-day postoperatively; 90-day = 90-day postoperatively; 180-day = 180-day postoperatively. Comparison with match HCs: \**P* < 0.05, \*\*\**P* < 0.001; comparison with Pre-op: ^#^*P* < 0.05, ^##^*P* < 0.01, ^##^*P* < 0.001.).

**Table 1.** Demographics for ET patients and their matched health controls.

| Variables | ET patients | health controls |
| --- | --- | --- |
| n | 31 | 31 |
| Age, y, mean (SD) | 62.16 (10.88) | 62.20 (10.86) |
| Sex, M/F | 21/10 | 21/10 |
| Education level, y, mean (SD) | 18.00 (3.35) | 18.000 (4.35) |
| Height, cm, mean (SD) | 169.55 (7.60) | 169.82 (7.40) |
| Weight, kg, mean (SD) | 72.52 (11.72) | 72.60 (11.83) |
| Duration, mean (SD) | 18.03 (10.35) | / |
| ET family history, Y/N | 22/9 | / |
| SDR, mean (SD) | 0.50 (0.11) | / |
| PHQ-9, mean (SD) | 2.06 (2.11) | 2.09 (2.09) |
| MMSE/30 | 28.48 (1. 23) | 29 (1.24) |

Note: MMSE = Mini-Mental State Examination; SDR = skull density ratio; PHQ-9 = The Patient Health Questionnaire (PHQ-9). All values are group means (with SD), except for constrained variables (MMSE), which are shown as median (interquartile range). Measures are uncorrected for any covariables.

**Table 2.** Tremor symptoms relieve in all MRgFUS patients.

| Clinical ratings/timepoint | Statics (mean ± SD)<sup>*</sup> | β-value<sup>&</sup> | SE<sup>&</sup> | $t$-value<sup>&</sup> | $P$-value<sup>&</sup> |
| --- | --- | --- | --- | --- | --- |
| **Hand tremor score** | | | | | $P_{for\ trend} < 0.001^{\#}$ |
| Preoperative state | 21.23 ± 4.23 | | | | |
| Postoperative 30-day | 3.48 ± 3.62 | -0.589 | 0.022 | -26.354 | < 0.001 |
| Postoperative 90-day | 5.27 ± 5.66 | -0.177 | 0.010 | -18.185 | < 0.001 |
| Postoperative 180-day | 5.27 ± 5.86 | -0.088 | 0.004 | -19.653 | < 0.001 |
| **CRST-A** | | | | | $P_{for\ trend} < 0.001^{\#}$ |
| Preoperative state | 15.90 ± 4.49 | | | | |
| Postoperative 30-day | 6.96 ± 4.34 | -0.289 | 0.021 | -13.571 | < 0.001 |
| Postoperative 90-day | 7.23 ± 4.98 | -0.098 | 0.008 | -12.092 | < 0.001 |
| Postoperative 180-day | 7.53 ± 5.88 | -0.046 | 0.005 | -9.923 | < 0.001 |
| **CRST-B** | | | | | $P_{for\ trend} < 0.001^{\#}$ |
| Preoperative state | 24.35 ± 6.84 | | | | |
| Postoperative 30-day | 10.04 ± 7.00 | -0.481 | 0.034 | -14.349 | < 0.001 |
| Postoperative 90-day | 12.50 ± 7.87 | -0.133 | 0.011 | -11.986 | < 0.001 |
| Postoperative 180-day | 11.33 ± 6.88 | -0.071 | 0.005 | -13.977 | < 0.001 |
| **CRST-C** | | | | | $P_{for\ trend} < 0.001^{\#}$ |
| Preoperative state | 17.35 ± 3.93 | | | | |
| Postoperative 30-day | 2.22 ± 3.58 | -0.513 | 0.029 | -17.865 | < 0.001 |
| Postoperative 90-day | 3.55 ± 5.36 | -0.156 | 0.012 | -13.416 | < 0.001 |
| Postoperative 180-day | 3.87 ± 5.68 | -0.074 | 0.005 | -14.654 | < 0.001 |
| **CRST-total** | | | | | $P_{for\ trend} < 0.001^{\#}$ |
| Preoperative state | 57.61 ± 11.54 | | | | |
| Postoperative 30-day | 19.22 ± 12.62 | -1.291 | 0.062 | -20.890 | < 0.001 |
| Postoperative 90-day | 22.27 ± 16.54 | -0.390 | 0.021 | -18.394 | < 0.001 |
| Postoperative 180-day | 22.73 ± 16.78 | -0.191 | 0.011 | -17.499 | < 0.001 |

Note: Among the variables, $\beta$ is the regression coefficient; SE is the standard error; $t$ is the difference statistic; $P$ value is the significance value; $P_{for\ trend}$ represents statistical significance in the LMM model.

### 3.1 Global and regional brain networks presented different postoperative immediate and longitudinal alterations after MRgFUS thalamotomy

To study the longitudinal alterations of functional connectivities after MRgFUS thalamotomy, alteration distribution maps of global functional connectivities were established compared to the match HCs by paired *t*-test. The alterations of whole-brain functional connectivities presented an approximately normal distribution across all the postoperative timepoints (Figure 1B). The alteration distribution shifted positively after MRgFUS thalamotomy and reached peak at postoperative 7-day, then began to recover and almost returned to Pre-op state at postoperative 180-day (Figure 1B). The hemisphere lateralization of global functional network was also analyzed and found no significant change (Figure 1C).

To better identify long-term functional features after MRgFUS thalamotomy, network-based statistics were performed based on the global functional network between postoperative 180-day and Pre-op timepoint. It revealed an enhanced functional subnetwork in ET patients at 180-day postoperatively (*t* = 4), comprising 186 edges and 124 nodes (*P* = 0.019, Figure 1D). No other significant subnetwork was identified in the opposite direction. The alteration distribution of this subnetwork shifted positively after MRgFUS thalamotomy and kept a large positive gap from Pre-op state at 180-day postoperatively (Figure 1E). The pre-operative hemisphere lateralization of MRgFUS thalamotomy subnetwork was much higher than the matched HC (*P* < 0.001, Figure 1F), and presented a remarkable recovery after MRgFUS thalamotomy (LMM: *P*_for trend_ < 0.001, Figure 1F).

Graph-theory network analysis was applied to both whole-brain functional network and MRgFUS thalamotomy subnetwork. Although there were short-term fluctuations (network degree, global efficiency and network transitivity: *P* < 0.05 at 7-day *vs.* Pre-op), it was found that network degree, global efficiency and network transitivity did not present significant alterations compared with the preoperative timepoint (all *P* > 0.05 at 180-day *vs.* Pre-op) (Figure G). Within this MRgFUS thalamotomy subnetwork, network degree, global efficiency and transitivity were significantly increased in ET patients after MRgFUS thalamotomy compared to the pre-operative timepoint (all *P* < 0.05 at different postoperative timepoints *vs.* Pre-op; LMM: *P*_for trend_ < 0.001, Figure 2H). The results of graph-theory analysis were robust when controlling for different connectivity matrix thresholding (15-30%)(Figure S1).

**Figure 2.**
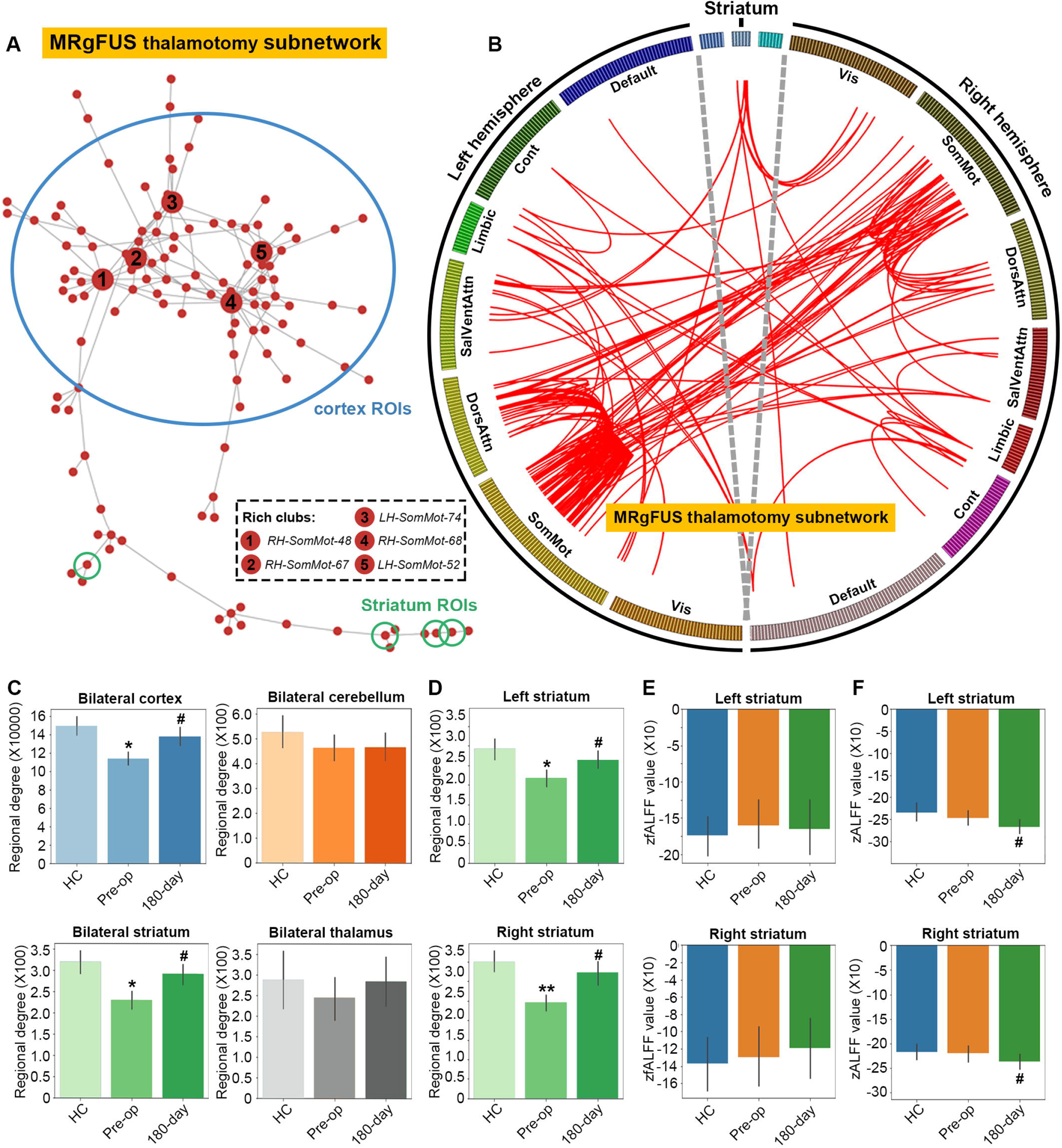
MRgFUS thalamotomy promoted activation of corticostriatal connectivity in ET patients. **(A and B)** The pyechart and circo presentation of MRgFUS thalamotomy subnetwork. **(C and D)** The regional degree of each anatomical region. **(E and F)** The zfALFF and zALFF results of striatum. (Pre-op = pre-operative; 1-day = 1-day postoperatively; 7-day = 7-day postoperatively; 30-day = 30-day postoperatively; 90-day = 90-day postoperatively; 365-day = 365-day postoperatively. Comparison with match HCs: \**P* < 0.05, \*\**P* < 0.01; comparison with Pre-op: ^#^*P* < 0.05.).

### 3.2 MRgFUS thalamotomy promoted activation of corticostriatal connectivity in ET patients

Then, we examined the functional characteristics of MRgFUS thalamotomy subnetwork. The subnetwork present significant corticostriatal feature with multiple functional connectivities with striatum and cortex, including *LH-Limbic-Striatum* to *LH-Vis-55*/*LH-Vis-63*/*LH-Vis-69*/*LH-Default-pCunPCC4*, *RH-SalVentAttn-Striatum* to *LH-Limbic-TempPole13*/*RH-SalVentAttn-FrOperIns12*/*RH-SalVentAttn-FrOperIns16*, *RH-Limbic-Striatum* to *LH-Vis-55*, *RH-Cont-Striatum* to *LH-Vis-55*/*RH-SomMot-6* (Figure 2A). Our further analysis of rich clubs suggested that *RH-SomMot-48*, *RH-SomMot-67*, *RH-SomMot-68*, *LH-SomMot-52*, *LH-SomMot-74* sustain the highest nodal degree within subnetwork (Figure 2A). The cicos scheme based on Yeo parcellation was built, and a notable feature of the corticostriatal subnetwork was the large number of functional connectivities in sensorimotor network (SMN)(Figure 2B).

In order to further analyze the alteration in anatomical regions, the regional functional strength features were calculated. There were no significant differences of regional degree for bilateral cerebellum, and thalamus (both *P* 0.05, Figure 2C). while regional degree in bilateral cortex and striatum was significantly decreased in the ET patients compared with matched HCs (bilateral cortex: Pre-op *vs*. HC: *P* = 0.010; bilateral striatum: Pre-op *vs*. HC: *P* = 0.012), which were rescued after MRgFUS thalamotomy (bilateral cortex: 180-day *vs*. Pre-op: *P* = 0.044; bilateral striatum: 180-day *vs*. Pre-op: *P* = 0.030)(Figure 2C). Then left and right striatum were separated, and it was found that regional degree of left and right striatum decreased significantly in ET patients compared with matched HCs (left striatum: Pre-op *vs*. HC: *P* = 0.037; right striatum: Pre-op *vs*. HC: *P* = 0.004), which presented an obvious increased after MRgFUS thalamotomy (left striatum: 180-day *vs*. Pre-op: *P* = 0.048; right striatum: 180-day *vs*. Pre-op: *P* = 0.047)(Figure 2D). To further illustrate the alteration in the striatum, zfALFF and zALFF were calculated respectively. The results found that zfALFF did not present significant alteration (Figure 2E), while zALFF showed obvious increase after MRgFUS thalamotomy (left striatum: 180-day *vs*. Pre-op: *P* = 0.019; right striatum: 180-day *vs*. Pre-op: *P* = 0.040)(Figure 2F).

### 3.3 Regional network features reflected tremor progression after MRgFUS thalamotomy

PLS, a multivariate statistical method, was applied to test whether functional connectivities of corticostriatal subnetwork could provide a common explanation for symptom relieve of MRgFUS thalamotomy. It was found that the PLS main component significantly accounted for 33.68%, 38.57%, 23.02%, 34.20% and 34.16% of the total variances of hand tremor score, CRST-A score, right CRST-B score, CRST-C score and CRST-total score (*P* = 0.037, 0.003, 0.036, 0.025 and 0.027 respectively)(Table 3). The predicted outcomes based on the PLS main component weights were significantly correlated with tremor ratings (hand tremor score: *r* = 0.58, *P* = 4.61 × 10^-3^; CRST-A: *r* = 0.62, *P* = 7.49 × 10^-13^; right CRST-B: *r* = 0.48, *P* = 1.48 × 10^-7^; CRST-C: *r* = 0.59, *P* = 2.93 × 10^-11^; CRST-total: *r* = 0.58, *P* = 3.13 × 10^-11^)(Figure 3A). Moreover, we investigated the correlations between functional metrics and tremor ratings of ET patients subjected with MRgFUS thalamotomy. The network degree, global efficiency, network transitivity of the corticostriatal subnetwork significantly correlated with hand tremor score (network degree: *r* = −0.35, *P* = 2.47 × 10^-4^; global efficiency: *r* = −0.21, *P* = 0.033; network transitivity: *r* = −0.34, *P* = 2.77 × 10^-4^)(Figure 3B). The hemisphere lateralization of corticostriatal subnetwork was also analyzed and found no significant correlation (Figure 3C).

**Figure 3.**
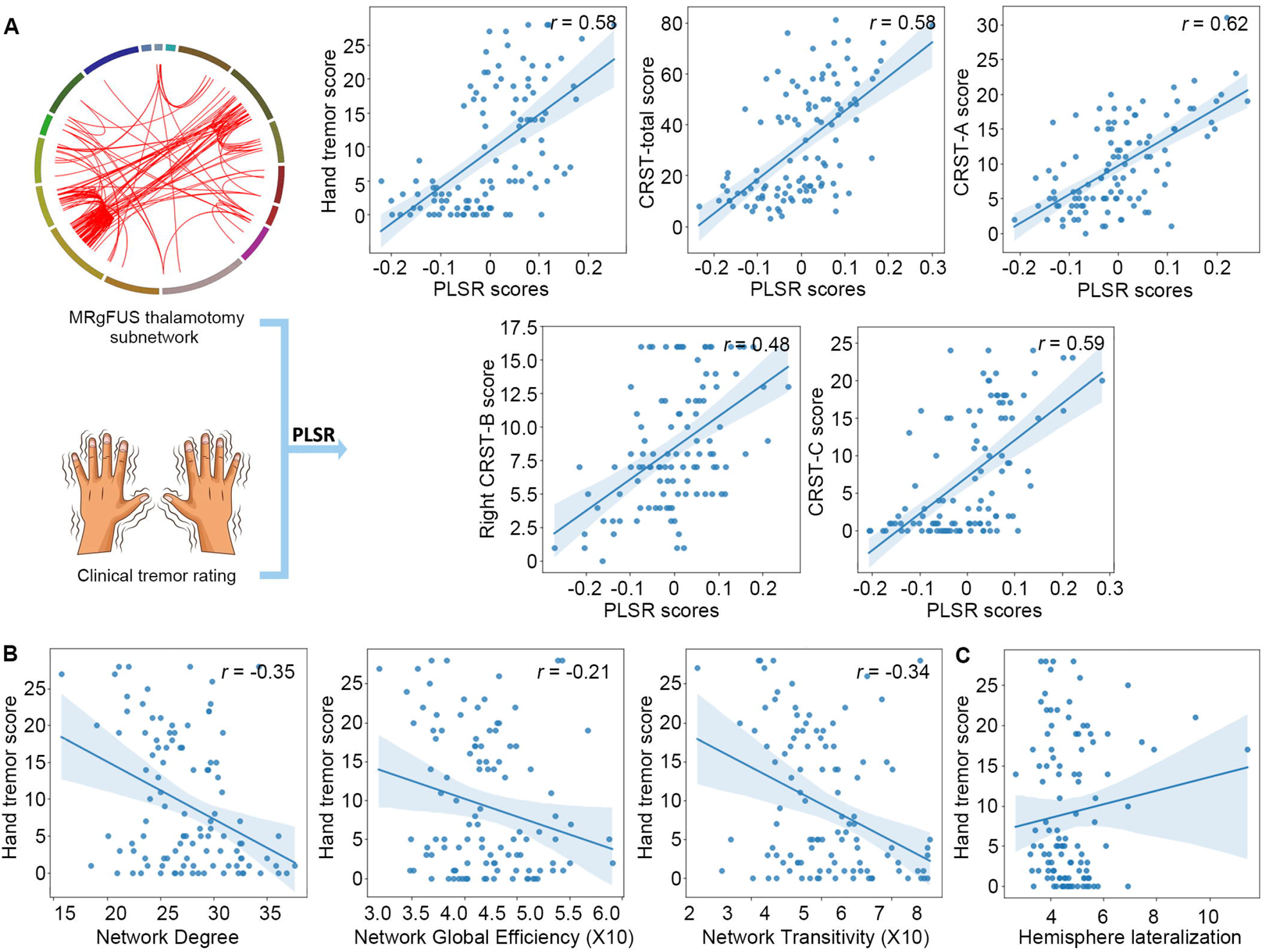
Regional network features reflected tremor progression after MRgFUS thalamotomy. **(A)** Overview of PLS regression. The Pearson correlation analysis for the predicted outcomes based on the PLS main component weights at 365-day postoperatively. **(B and C)** Correlation between graph-theory metrics and hemisphere lateralization of corticostriatal subnetwork and hand tremor scores

**Table 3.**
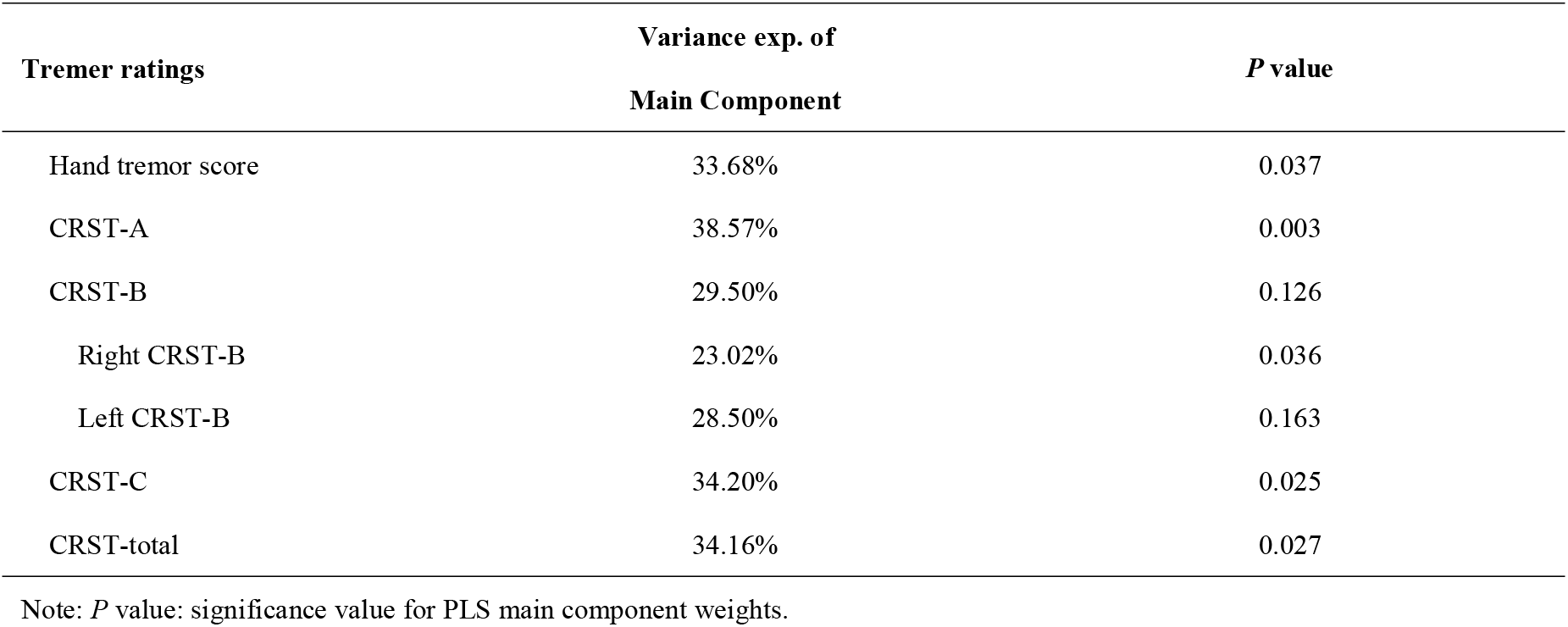
PLS regression for ET tremor ratings.

We utilized supervised statistical learning to test whether pre-operative functional connectivities can provide a prediction for tremor relieves at 180-day postoperatively. Using three-fold cross-validations, there was no significant performance for hand tremor score or any tremor assessments. Furthermore, pre-operative functional metrics (network degree, global efficiency, network transitivity) of corticostriatal subnetwork were also applied in linear regression for tremor prediction with adjustment of sex and age. Post hoc analysis found only network transitivity could serve as a reliable biomarker for hand tremor score control prediction (network degree: *P* 0.05, global efficiency: *P* 0.05, network transitivity: β = 2.94, *P* = 0.03).

## 4. Discussion

In the present study, we fully profiling the functional alteration of MRgFUS thalamotomy in medication-refractory ET patients. An enhanced corticostriatal subnetwork correlated with tremor improvement was identified starting from the first day after thalamotomy, while there was an obvious and immediate fluctuation within one week postoperatively in the global network but stabilized after six months. This regional subnetwork presented well-defined corticostriatal features, and the functional communication within the striatum were also significantly recovered post MRgFUS thalamotomy. Not only the functional features of this corticostriatal subnetwork (connectivity features and graph-theory metrics) better reflected the tremor symptoms in ET patients, but also network transitivity of subnetwork could serve as a reliable biomarker for hand tremor score control prediction.

The understanding of immediate and longitudinal alterations of functional networks after thalamotomy has always been valued by clinicians. From the broad perspective, in our present study, the single Vim lesion did not cause significant global functional alteration. Interestingly, there was indeed an obvious fluctuation within postoperative one week, not only reflected in altered functional connectivity distribution but also in the global function network metrics. However, in the previous studies of Jang et al. [4], they found that the Vim ablation induced a transient decrease of average node strength in the whole brain network, which then increased after seven days postoperatively. This difference, on the one hand, may be due to local brain edema, associated distal nucleus damage, and “transient connectional and connectomal diaschisis effects due to focal lesion” as they reported; on the other hand, it may also be due to the small sample size (n = 8) and gender bias (M: F = 7: 1) in their study [4]. Some recent studies suggested that low-frequency focused ultrasound, low-energy ultrasound could cause transient blood-brain barrier opening and neuromodulation effects [30–32], which may be a concrete manifestation for the immediate effects in our high-frequency ultrasound therapy. In any case, these all suggested that it may not be appropriate to perform functional studies in the early postoperative period of MRgFUS thalamotomy.

Contrary to the global functional network stability, there was significant enhanced, long-term alteration in corticostriatal subnetwork starting from the first day after MRgFUS thalamotomy. As expected, the functional connectivities and metrics features in the subnetwork not only reflected the improvement of clinical tremor symptoms, but also were closely related to SMN. However, it was clearly observed that a significant perturbated connectivities among the cortical rather than subcortical regions, which is consistent with our recent analysis of functional gradient post MRgFUS thalamotomy [5]. Vim lesion did not result in significant gradient alteration in subcortical nuclei, including injured thalamus, but extensive reorganization of the cortical gradients [5]. Our previous study also found no significant change in the neural spontaneous activity of the Vim lesion site, while decreased in the left occipital cortex after MRgFUS thalamotomy [33]. The rs-fMRI study from Jang et al. and Mario et al. indicated a significant perturbation in the cortical network rather than the thalamus as well [4, 34]. This may be a manifestation of neuro-dynamics compensation or brain structure remodeling; another possible explanation for this phenomenon is that our 3T MRI-based imaging scans have relatively limited capabilities for subcortical tissue, restricting us to capture more subtle changes. It may be helpful to perform functional imaging studies at higher field strengths such as 7T. From another perspective, it also suggested that cortical activity could accurately reflect the subcortical neuro-regulation, and these robust cortical alterations were prone to be detected by other techniques (such as electroencephalogram, magnetoencephalography, etc). Other transcranial stimulation tools, such as transcranial magnetic stimulation or transcranial electrical stimulation, may be helpful to further consolidate the therapeutic effect of MRgFUS thalamotomy, which needs further exploration.

In present study, it was found that MRgFUS thalamotomy mainly regulated the functional activities of corticostriatal connectivity. Although the pathophysiology of tremor has not been fully understood to date, the results of surgical, neurophysiological, and postmortem studies indicate that cerebellum, red nucleus, thalamus, and cerebral cortex (known as dentato-rubro-thalamic tract or cerebello-thalamo-cortical circuits) are involved in ET pathomechanism [35], which is often compared to the striatal circuit (known as striatal-thalamo-cortical circuits) in PD pathomechanism [35]. Vim is a key feedback regulator in internal thalamus, which receives input from the cerebellum via the dentato-rubro-thalamic tract and also serves as a causal flow hub for striatal-thalamo-cortical circuits [36]. Therefore, thalamotomy is thought to be effective for both kinetic tremor of ET and resting tremor of PD [37]. However, compared with the limited alteration in the cerebellum, our postoperative study found that MRgFUS thalamotomy had the most significant impacts on the corticostriatal connectivity in ET patients. This suggests the different intervention effect of Vim on cerebello-thalamo-cortical and striatal-thalamo-cortical circuits. It is worthy for us to further better new therapeutic targets suitable for dentato-rubro-thalamic tract in ET pathogenesis.

Several methodological considerations should be contemplated when interpreting the results of this study. This study also lacks data on the longitudinal changes in healthy control groups. A small number of patients with extreme tremor may lead to low statistical power considering subjects’ heterogeneity. Replication in larger cohorts as well as longitudinal assessment at more time points following MRgFUS thalamotomy would provide further insights.

## Data Availability

The conditions of our ethics approval do not permit public archiving of raw data. All code and statistical image data are available at https://github.com/louxin-lab.

## Author contributions

Conceptualization: XL, LSP, JJL, XPK; Methodology: JJL, XPK, DKZ, JXH, HXL; Investigation: DKZ, JXH, XPK; Visualization: JJL, DKZ; Supervision: JJL, LSP, XL; Writing-original draft: JJL, XPK; Writing-review & editing: LSP, XL.

## Acknowledgments

This research was supported by National Natural Science Foundation of China 82151309 and 81825012 to XL as well as China Postdoctoral Science Foundation 2022T150788 to JJL.

