## Supplementary material for "Profiling functional networks identify activation of corticostriatal connectivity in ET patient after MRgFUS thalamotomy": Figure S1

### Table S1. Atlas annotation

**Table S1 Atlas annotation**

| ROI Label | ROI Name | Yeo community | Atlas source |
|---|---|---|---|
| 1 | 7Networks_LH_Vis_1 | Vis | Schaefer |
| 2 | 7Networks_LH_Vis_2 | Vis | Schaefer |
| 3 | 7Networks_LH_Vis_3 | Vis | Schaefer |
| 4 | 7Networks_LH_Vis_4 | Vis | Schaefer |
| 5 | 7Networks_LH_Vis_5 | Vis | Schaefer |
| 6 | 7Networks_LH_Vis_6 | Vis | Schaefer |
| 7 | 7Networks_LH_Vis_7 | Vis | Schaefer |
| 8 | 7Networks_LH_Vis_8 | Vis | Schaefer |
| 9 | 7Networks_LH_Vis_9 | Vis | Schaefer |
| 10 | 7Networks_LH_Vis_10 | Vis | Schaefer |
| 11 | 7Networks_LH_Vis_11 | Vis | Schaefer |
| 12 | 7Networks_LH_Vis_12 | Vis | Schaefer |
| 13 | 7Networks_LH_Vis_13 | Vis | Schaefer |
| 14 | 7Networks_LH_Vis_14 | Vis | Schaefer |
| 15 | 7Networks_LH_Vis_15 | Vis | Schaefer |
| 16 | 7Networks_LH_Vis_16 | Vis | Schaefer |
| 17 | 7Networks_LH_Vis_17 | Vis | Schaefer |
| 18 | 7Networks_LH_Vis_18 | Vis | Schaefer |
| 19 | 7Networks_LH_Vis_19 | Vis | Schaefer |
| 20 | 7Networks_LH_Vis_20 | Vis | Schaefer |
| 21 | 7Networks_LH_Vis_21 | Vis | Schaefer |
| 22 | 7Networks_LH_Vis_22 | Vis | Schaefer |
| 23 | 7Networks_LH_Vis_23 | Vis | Schaefer |
| 24 | 7Networks_LH_Vis_24 | Vis | Schaefer |
| 25 | 7Networks_LH_Vis_25 | Vis | Schaefer |
| 26 | 7Networks_LH_Vis_26 | Vis | Schaefer |
| 27 | 7Networks_LH_Vis_27 | Vis | Schaefer |
| 28 | 7Networks_LH_Vis_28 | Vis | Schaefer |
| 29 | 7Networks_LH_Vis_29 | Vis | Schaefer |
| 30 | 7Networks_LH_Vis_30 | Vis | Schaefer |
| 31 | 7Networks_LH_Vis_31 | Vis | Schaefer |
| 32 | 7Networks_LH_Vis_32 | Vis | Schaefer |
| 33 | 7Networks_LH_Vis_33 | Vis | Schaefer |
| 34 | 7Networks_LH_Vis_34 | Vis | Schaefer |
| 35 | 7Networks_LH_Vis_35 | Vis | Schaefer |
| 36 | 7Networks_LH_Vis_36 | Vis | Schaefer |
| 37 | 7Networks_LH_Vis_37 | Vis | Schaefer |
| 38 | 7Networks_LH_Vis_38 | Vis | Schaefer |
| 39 | 7Networks_LH_Vis_39 | Vis | Schaefer |
| 40 | 7Networks_LH_Vis_40 | Vis | Schaefer |
| 41 | 7Networks_LH_Vis_41 | Vis | Schaefer |

| | | | |
|---|---|---|---|
| 42 | 7Networks_LH_Vis_42 | Vis | Schaefer |
| 43 | 7Networks_LH_Vis_43 | Vis | Schaefer |
| 44 | 7Networks_LH_Vis_44 | Vis | Schaefer |
| 45 | 7Networks_LH_Vis_45 | Vis | Schaefer |
| 46 | 7Networks_LH_Vis_46 | Vis | Schaefer |
| 47 | 7Networks_LH_Vis_47 | Vis | Schaefer |
| 48 | 7Networks_LH_Vis_48 | Vis | Schaefer |
| 49 | 7Networks_LH_Vis_49 | Vis | Schaefer |
| 50 | 7Networks_LH_Vis_50 | Vis | Schaefer |
| 51 | 7Networks_LH_Vis_51 | Vis | Schaefer |
| 52 | 7Networks_LH_Vis_52 | Vis | Schaefer |
| 53 | 7Networks_LH_Vis_53 | Vis | Schaefer |
| 54 | 7Networks_LH_Vis_54 | Vis | Schaefer |
| 55 | 7Networks_LH_Vis_55 | Vis | Schaefer |
| 56 | 7Networks_LH_Vis_56 | Vis | Schaefer |
| 57 | 7Networks_LH_Vis_57 | Vis | Schaefer |
| 58 | 7Networks_LH_Vis_58 | Vis | Schaefer |
| 59 | 7Networks_LH_Vis_59 | Vis | Schaefer |
| 60 | 7Networks_LH_Vis_60 | Vis | Schaefer |
| 61 | 7Networks_LH_Vis_61 | Vis | Schaefer |
| 62 | 7Networks_LH_Vis_62 | Vis | Schaefer |
| 63 | 7Networks_LH_Vis_63 | Vis | Schaefer |
| 64 | 7Networks_LH_Vis_64 | Vis | Schaefer |
| 65 | 7Networks_LH_Vis_65 | Vis | Schaefer |
| 66 | 7Networks_LH_Vis_66 | Vis | Schaefer |
| 67 | 7Networks_LH_Vis_67 | Vis | Schaefer |
| 68 | 7Networks_LH_Vis_68 | Vis | Schaefer |
| 69 | 7Networks_LH_Vis_69 | Vis | Schaefer |
| 70 | 7Networks_LH_Vis_70 | Vis | Schaefer |
| 71 | 7Networks_LH_Vis_71 | Vis | Schaefer |
| 72 | 7Networks_LH_Vis_72 | Vis | Schaefer |
| 73 | 7Networks_LH_Vis_73 | Vis | Schaefer |
| 74 | 7Networks_LH_Vis_74 | Vis | Schaefer |
| 75 | 7Networks_LH_Vis_75 | Vis | Schaefer |
| 76 | 7Networks_LH_Vis_76 | Vis | Schaefer |
| 77 | 7Networks_LH_Vis_77 | Vis | Schaefer |
| 78 | 7Networks_LH_Vis_78 | Vis | Schaefer |
| 79 | 7Networks_LH_Vis_79 | Vis | Schaefer |
| 80 | 7Networks_LH_Vis_80 | Vis | Schaefer |
| 81 | 7Networks_LH_Vis_81 | Vis | Schaefer |
| 82 | 7Networks_LH_SomMot_1 | SomMot | Schaefer |
| 83 | 7Networks_LH_SomMot_2 | SomMot | Schaefer |
| 84 | 7Networks_LH_SomMot_3 | SomMot | Schaefer |
| 85 | 7Networks_LH_SomMot_4 | SomMot | Schaefer |
| 86 | 7Networks_LH_SomMot_5 | SomMot | Schaefer |

| | | | |
|---|---|---|---|
| 87 | 7Networks_LH_SomMot_6 | SomMot | Schaefer |
| 88 | 7Networks_LH_SomMot_7 | SomMot | Schaefer |
| 89 | 7Networks_LH_SomMot_8 | SomMot | Schaefer |
| 90 | 7Networks_LH_SomMot_9 | SomMot | Schaefer |
| 91 | 7Networks_LH_SomMot_10 | SomMot | Schaefer |
| 92 | 7Networks_LH_SomMot_11 | SomMot | Schaefer |
| 93 | 7Networks_LH_SomMot_12 | SomMot | Schaefer |
| 94 | 7Networks_LH_SomMot_13 | SomMot | Schaefer |
| 95 | 7Networks_LH_SomMot_14 | SomMot | Schaefer |
| 96 | 7Networks_LH_SomMot_15 | SomMot | Schaefer |
| 97 | 7Networks_LH_SomMot_16 | SomMot | Schaefer |
| 98 | 7Networks_LH_SomMot_17 | SomMot | Schaefer |
| 99 | 7Networks_LH_SomMot_18 | SomMot | Schaefer |
| 100 | 7Networks_LH_SomMot_19 | SomMot | Schaefer |
| 101 | 7Networks_LH_SomMot_20 | SomMot | Schaefer |
| 102 | 7Networks_LH_SomMot_21 | SomMot | Schaefer |
| 103 | 7Networks_LH_SomMot_22 | SomMot | Schaefer |
| 104 | 7Networks_LH_SomMot_23 | SomMot | Schaefer |
| 105 | 7Networks_LH_SomMot_24 | SomMot | Schaefer |
| 106 | 7Networks_LH_SomMot_25 | SomMot | Schaefer |
| 107 | 7Networks_LH_SomMot_26 | SomMot | Schaefer |
| 108 | 7Networks_LH_SomMot_27 | SomMot | Schaefer |
| 109 | 7Networks_LH_SomMot_28 | SomMot | Schaefer |
| 110 | 7Networks_LH_SomMot_29 | SomMot | Schaefer |
| 111 | 7Networks_LH_SomMot_30 | SomMot | Schaefer |
| 112 | 7Networks_LH_SomMot_31 | SomMot | Schaefer |
| 113 | 7Networks_LH_SomMot_32 | SomMot | Schaefer |
| 114 | 7Networks_LH_SomMot_33 | SomMot | Schaefer |
| 115 | 7Networks_LH_SomMot_34 | SomMot | Schaefer |
| 116 | 7Networks_LH_SomMot_35 | SomMot | Schaefer |
| 117 | 7Networks_LH_SomMot_36 | SomMot | Schaefer |
| 118 | 7Networks_LH_SomMot_37 | SomMot | Schaefer |
| 119 | 7Networks_LH_SomMot_38 | SomMot | Schaefer |
| 120 | 7Networks_LH_SomMot_39 | SomMot | Schaefer |
| 121 | 7Networks_LH_SomMot_40 | SomMot | Schaefer |
| 122 | 7Networks_LH_SomMot_41 | SomMot | Schaefer |
| 123 | 7Networks_LH_SomMot_42 | SomMot | Schaefer |
| 124 | 7Networks_LH_SomMot_43 | SomMot | Schaefer |
| 125 | 7Networks_LH_SomMot_44 | SomMot | Schaefer |
| 126 | 7Networks_LH_SomMot_45 | SomMot | Schaefer |
| 127 | 7Networks_LH_SomMot_46 | SomMot | Schaefer |
| 128 | 7Networks_LH_SomMot_47 | SomMot | Schaefer |
| 129 | 7Networks_LH_SomMot_48 | SomMot | Schaefer |
| 130 | 7Networks_LH_SomMot_49 | SomMot | Schaefer |
| 131 | 7Networks_LH_SomMot_50 | SomMot | Schaefer |

| | | | |
|---|---|---|---|
| 132 | 7Networks_LH_SomMot_51 | SomMot | Schaefer |
| 133 | 7Networks_LH_SomMot_52 | SomMot | Schaefer |
| 134 | 7Networks_LH_SomMot_53 | SomMot | Schaefer |
| 135 | 7Networks_LH_SomMot_54 | SomMot | Schaefer |
| 136 | 7Networks_LH_SomMot_55 | SomMot | Schaefer |
| 137 | 7Networks_LH_SomMot_56 | SomMot | Schaefer |
| 138 | 7Networks_LH_SomMot_57 | SomMot | Schaefer |
| 139 | 7Networks_LH_SomMot_58 | SomMot | Schaefer |
| 140 | 7Networks_LH_SomMot_59 | SomMot | Schaefer |
| 141 | 7Networks_LH_SomMot_60 | SomMot | Schaefer |
| 142 | 7Networks_LH_SomMot_61 | SomMot | Schaefer |
| 143 | 7Networks_LH_SomMot_62 | SomMot | Schaefer |
| 144 | 7Networks_LH_SomMot_63 | SomMot | Schaefer |
| 145 | 7Networks_LH_SomMot_64 | SomMot | Schaefer |
| 146 | 7Networks_LH_SomMot_65 | SomMot | Schaefer |
| 147 | 7Networks_LH_SomMot_66 | SomMot | Schaefer |
| 148 | 7Networks_LH_SomMot_67 | SomMot | Schaefer |
| 149 | 7Networks_LH_SomMot_68 | SomMot | Schaefer |
| 150 | 7Networks_LH_SomMot_69 | SomMot | Schaefer |
| 151 | 7Networks_LH_SomMot_70 | SomMot | Schaefer |
| 152 | 7Networks_LH_SomMot_71 | SomMot | Schaefer |
| 153 | 7Networks_LH_SomMot_72 | SomMot | Schaefer |
| 154 | 7Networks_LH_SomMot_73 | SomMot | Schaefer |
| 155 | 7Networks_LH_SomMot_74 | SomMot | Schaefer |
| 156 | 7Networks_LH_SomMot_75 | SomMot | Schaefer |
| 157 | 7Networks_LH_SomMot_76 | SomMot | Schaefer |
| 158 | 7Networks_LH_SomMot_77 | SomMot | Schaefer |
| 159 | 7Networks_LH_SomMot_78 | SomMot | Schaefer |
| 160 | 7Networks_LH_SomMot_79 | SomMot | Schaefer |
| 161 | 7Networks_LH_SomMot_80 | SomMot | Schaefer |
| 162 | 7Networks_LH_SomMot_81 | SomMot | Schaefer |
| 163 | 7Networks_LH_SomMot_82 | SomMot | Schaefer |
| 164 | 7Networks_LH_SomMot_83 | SomMot | Schaefer |
| 165 | 7Networks_LH_SomMot_84 | SomMot | Schaefer |
| 166 | 7Networks_LH_SomMot_85 | SomMot | Schaefer |
| 167 | 7Networks_LH_SomMot_86 | SomMot | Schaefer |
| 168 | 7Networks_LH_SomMot_87 | SomMot | Schaefer |
| 169 | 7Networks_LH_SomMot_88 | SomMot | Schaefer |
| 170 | 7Networks_LH_SomMot_89 | SomMot | Schaefer |
| 171 | 7Networks_LH_SomMot_90 | SomMot | Schaefer |
| 172 | 7Networks_LH_SomMot_91 | SomMot | Schaefer |
| 173 | 7Networks_LH_DorsAttn_Post_1 | SalVentAt | Schaefer |
| 174 | 7Networks_LH_DorsAttn_Post_2 | SalVentAt | Schaefer |
| 175 | 7Networks_LH_DorsAttn_Post_3 | SalVentAt | Schaefer |
| 176 | 7Networks_LH_DorsAttn_Post_4 | SalVentAt | Schaefer |

| | | | |
|---|---|---|---|
| 177 | 7Networks_LH_DorsAttn_Post_5 | SalVentAt | Schaefer |
| 178 | 7Networks_LH_DorsAttn_Post_6 | SalVentAt | Schaefer |
| 179 | 7Networks_LH_DorsAttn_Post_7 | SalVentAt | Schaefer |
| 180 | 7Networks_LH_DorsAttn_Post_8 | SalVentAt | Schaefer |
| 181 | 7Networks_LH_DorsAttn_Post_9 | SalVentAt | Schaefer |
| 182 | 7Networks_LH_DorsAttn_Post_10 | SalVentAt | Schaefer |
| 183 | 7Networks_LH_DorsAttn_Post_11 | SalVentAt | Schaefer |
| 184 | 7Networks_LH_DorsAttn_Post_12 | SalVentAt | Schaefer |
| 185 | 7Networks_LH_DorsAttn_Post_13 | SalVentAt | Schaefer |
| 186 | 7Networks_LH_DorsAttn_Post_14 | SalVentAt | Schaefer |
| 187 | 7Networks_LH_DorsAttn_Post_15 | SalVentAt | Schaefer |
| 188 | 7Networks_LH_DorsAttn_Post_16 | SalVentAt | Schaefer |
| 189 | 7Networks_LH_DorsAttn_Post_17 | SalVentAt | Schaefer |
| 190 | 7Networks_LH_DorsAttn_Post_18 | SalVentAt | Schaefer |
| 191 | 7Networks_LH_DorsAttn_Post_19 | SalVentAt | Schaefer |
| 192 | 7Networks_LH_DorsAttn_Post_20 | SalVentAt | Schaefer |
| 193 | 7Networks_LH_DorsAttn_Post_21 | SalVentAt | Schaefer |
| 194 | 7Networks_LH_DorsAttn_Post_22 | SalVentAt | Schaefer |
| 195 | 7Networks_LH_DorsAttn_Post_23 | SalVentAt | Schaefer |
| 196 | 7Networks_LH_DorsAttn_Post_24 | SalVentAt | Schaefer |
| 197 | 7Networks_LH_DorsAttn_Post_25 | SalVentAt | Schaefer |
| 198 | 7Networks_LH_DorsAttn_Post_26 | SalVentAt | Schaefer |
| 199 | 7Networks_LH_DorsAttn_Post_27 | SalVentAt | Schaefer |
| 200 | 7Networks_LH_DorsAttn_Post_28 | SalVentAt | Schaefer |
| 201 | 7Networks_LH_DorsAttn_Post_29 | SalVentAt | Schaefer |
| 202 | 7Networks_LH_DorsAttn_Post_30 | SalVentAt | Schaefer |
| 203 | 7Networks_LH_DorsAttn_Post_31 | SalVentAt | Schaefer |
| 204 | 7Networks_LH_DorsAttn_Post_32 | SalVentAt | Schaefer |
| 205 | 7Networks_LH_DorsAttn_Post_33 | SalVentAt | Schaefer |
| 206 | 7Networks_LH_DorsAttn_Post_34 | SalVentAt | Schaefer |
| 207 | 7Networks_LH_DorsAttn_Post_35 | SalVentAt | Schaefer |
| 208 | 7Networks_LH_DorsAttn_Post_36 | SalVentAt | Schaefer |
| 209 | 7Networks_LH_DorsAttn_Post_37 | SalVentAt | Schaefer |
| 210 | 7Networks_LH_DorsAttn_Post_38 | SalVentAt | Schaefer |
| 211 | 7Networks_LH_DorsAttn_Post_39 | SalVentAt | Schaefer |
| 212 | 7Networks_LH_DorsAttn_Post_40 | SalVentAt | Schaefer |
| 213 | 7Networks_LH_DorsAttn_Post_41 | SalVentAt | Schaefer |
| 214 | 7Networks_LH_DorsAttn_Post_42 | SalVentAt | Schaefer |
| 215 | 7Networks_LH_DorsAttn_Post_43 | SalVentAt | Schaefer |
| 216 | 7Networks_LH_DorsAttn_Post_44 | SalVentAt | Schaefer |
| 217 | 7Networks_LH_DorsAttn_Post_45 | SalVentAt | Schaefer |
| 218 | 7Networks_LH_DorsAttn_Post_46 | SalVentAt | Schaefer |
| 219 | 7Networks_LH_DorsAttn_Post_47 | SalVentAt | Schaefer |
| 220 | 7Networks_LH_DorsAttn_Post_48 | SalVentAt | Schaefer |
| 221 | 7Networks_LH_DorsAttn_Post_49 | SalVentAt | Schaefer |

| | | | |
|---|---|---|---|
| 222 | 7Networks_LH_DorsAttn_Post_50 | SalVentAt | Schaefer |
| 223 | 7Networks_LH_DorsAttn_FEF_1 | SalVentAt | Schaefer |
| 224 | 7Networks_LH_DorsAttn_FEF_2 | SalVentAt | Schaefer |
| 225 | 7Networks_LH_DorsAttn_FEF_3 | SalVentAt | Schaefer |
| 226 | 7Networks_LH_DorsAttn_FEF_4 | SalVentAt | Schaefer |
| 227 | 7Networks_LH_DorsAttn_FEF_5 | SalVentAt | Schaefer |
| 228 | 7Networks_LH_DorsAttn_FEF_6 | SalVentAt | Schaefer |
| 229 | 7Networks_LH_DorsAttn_FEF_7 | SalVentAt | Schaefer |
| 230 | 7Networks_LH_DorsAttn_PrCv_1 | SalVentAt | Schaefer |
| 231 | 7Networks_LH_DorsAttn_PrCv_2 | SalVentAt | Schaefer |
| 232 | 7Networks_LH_DorsAttn_PrCv_3 | SalVentAt | Schaefer |
| 233 | 7Networks_LH_DorsAttn_PrCv_4 | SalVentAt | Schaefer |
| 234 | 7Networks_LH_SalVentAttn_ParOper_1 | DorsAttn | Schaefer |
| 235 | 7Networks_LH_SalVentAttn_ParOper_2 | DorsAttn | Schaefer |
| 236 | 7Networks_LH_SalVentAttn_ParOper_3 | DorsAttn | Schaefer |
| 237 | 7Networks_LH_SalVentAttn_ParOper_4 | DorsAttn | Schaefer |
| 238 | 7Networks_LH_SalVentAttn_ParOper_5 | DorsAttn | Schaefer |
| 239 | 7Networks_LH_SalVentAttn_ParOper_6 | DorsAttn | Schaefer |
| 240 | 7Networks_LH_SalVentAttn_ParOper_7 | DorsAttn | Schaefer |
| 241 | 7Networks_LH_SalVentAttn_ParOper_8 | DorsAttn | Schaefer |
| 242 | 7Networks_LH_SalVentAttn_ParOper_9 | DorsAttn | Schaefer |
| 243 | 7Networks_LH_SalVentAttn_TempOcc_1 | DorsAttn | Schaefer |
| 244 | 7Networks_LH_SalVentAttn_TempOcc_2 | DorsAttn | Schaefer |
| 245 | 7Networks_LH_SalVentAttn_FrOperIns_1 | DorsAttn | Schaefer |
| 246 | 7Networks_LH_SalVentAttn_FrOperIns_2 | DorsAttn | Schaefer |
| 247 | 7Networks_LH_SalVentAttn_FrOperIns_3 | DorsAttn | Schaefer |
| 248 | 7Networks_LH_SalVentAttn_FrOperIns_4 | DorsAttn | Schaefer |
| 249 | 7Networks_LH_SalVentAttn_FrOperIns_5 | DorsAttn | Schaefer |
| 250 | 7Networks_LH_SalVentAttn_FrOperIns_6 | DorsAttn | Schaefer |
| 251 | 7Networks_LH_SalVentAttn_FrOperIns_7 | DorsAttn | Schaefer |
| 252 | 7Networks_LH_SalVentAttn_FrOperIns_8 | DorsAttn | Schaefer |
| 253 | 7Networks_LH_SalVentAttn_FrOperIns_9 | DorsAttn | Schaefer |
| 254 | 7Networks_LH_SalVentAttn_FrOperIns_10 | DorsAttn | Schaefer |
| 255 | 7Networks_LH_SalVentAttn_FrOperIns_11 | DorsAttn | Schaefer |
| 256 | 7Networks_LH_SalVentAttn_FrOperIns_12 | DorsAttn | Schaefer |
| 257 | 7Networks_LH_SalVentAttn_FrOperIns_13 | DorsAttn | Schaefer |
| 258 | 7Networks_LH_SalVentAttn_FrOperIns_14 | DorsAttn | Schaefer |
| 259 | 7Networks_LH_SalVentAttn_FrOperIns_15 | DorsAttn | Schaefer |
| 260 | 7Networks_LH_SalVentAttn_FrOperIns_16 | DorsAttn | Schaefer |
| 261 | 7Networks_LH_SalVentAttn_FrOperIns_17 | DorsAttn | Schaefer |
| 262 | 7Networks_LH_SalVentAttn_FrOperIns_18 | DorsAttn | Schaefer |
| 263 | 7Networks_LH_SalVentAttn_FrOperIns_19 | DorsAttn | Schaefer |
| 264 | 7Networks_LH_SalVentAttn_FrOperIns_20 | DorsAttn | Schaefer |
| 265 | 7Networks_LH_SalVentAttn_FrOperIns_21 | DorsAttn | Schaefer |
| 266 | 7Networks_LH_SalVentAttn_FrOperIns_22 | DorsAttn | Schaefer |

| | | | |
|---|---|---|---|
| 267 | 7Networks_LH_SalVentAttn_FrOperIns_23 | DorsAttn | Schaefer |
| 268 | 7Networks_LH_SalVentAttn_PFCl_1 | DorsAttn | Schaefer |
| 269 | 7Networks_LH_SalVentAttn_PFCl_2 | DorsAttn | Schaefer |
| 270 | 7Networks_LH_SalVentAttn_Med_1 | DorsAttn | Schaefer |
| 271 | 7Networks_LH_SalVentAttn_Med_2 | DorsAttn | Schaefer |
| 272 | 7Networks_LH_SalVentAttn_Med_3 | DorsAttn | Schaefer |
| 273 | 7Networks_LH_SalVentAttn_Med_4 | DorsAttn | Schaefer |
| 274 | 7Networks_LH_SalVentAttn_Med_5 | DorsAttn | Schaefer |
| 275 | 7Networks_LH_SalVentAttn_Med_6 | DorsAttn | Schaefer |
| 276 | 7Networks_LH_SalVentAttn_Med_7 | DorsAttn | Schaefer |
| 277 | 7Networks_LH_SalVentAttn_Med_8 | DorsAttn | Schaefer |
| 278 | 7Networks_LH_SalVentAttn_Med_9 | DorsAttn | Schaefer |
| 279 | 7Networks_LH_SalVentAttn_Med_10 | DorsAttn | Schaefer |
| 280 | 7Networks_LH_SalVentAttn_Med_11 | DorsAttn | Schaefer |
| 281 | 7Networks_LH_SalVentAttn_Med_12 | DorsAttn | Schaefer |
| 282 | 7Networks_LH_SalVentAttn_Med_13 | DorsAttn | Schaefer |
| 283 | 7Networks_LH_SalVentAttn_Med_14 | DorsAttn | Schaefer |
| 284 | 7Networks_LH_SalVentAttn_Med_15 | DorsAttn | Schaefer |
| 285 | 7Networks_LH_SalVentAttn_Med_16 | DorsAttn | Schaefer |
| 286 | 7Networks_LH_SalVentAttn_Med_17 | DorsAttn | Schaefer |
| 287 | 7Networks_LH_SalVentAttn_Med_18 | DorsAttn | Schaefer |
| 288 | 7Networks_LH_SalVentAttn_Med_19 | DorsAttn | Schaefer |
| 289 | 7Networks_LH_Limbic_OFC_1 | Limbic | Schaefer |
| 290 | 7Networks_LH_Limbic_OFC_2 | Limbic | Schaefer |
| 291 | 7Networks_LH_Limbic_OFC_3 | Limbic | Schaefer |
| 292 | 7Networks_LH_Limbic_OFC_4 | Limbic | Schaefer |
| 293 | 7Networks_LH_Limbic_OFC_5 | Limbic | Schaefer |
| 294 | 7Networks_LH_Limbic_OFC_6 | Limbic | Schaefer |
| 295 | 7Networks_LH_Limbic_OFC_7 | Limbic | Schaefer |
| 296 | 7Networks_LH_Limbic_OFC_8 | Limbic | Schaefer |
| 297 | 7Networks_LH_Limbic_OFC_9 | Limbic | Schaefer |
| 298 | 7Networks_LH_Limbic_OFC_10 | Limbic | Schaefer |
| 299 | 7Networks_LH_Limbic_OFC_11 | Limbic | Schaefer |
| 300 | 7Networks_LH_Limbic_OFC_12 | Limbic | Schaefer |
| 301 | 7Networks_LH_Limbic_OFC_13 | Limbic | Schaefer |
| 302 | 7Networks_LH_Limbic_OFC_14 | Limbic | Schaefer |
| 303 | 7Networks_LH_Limbic_TempPole_1 | Limbic | Schaefer |
| 304 | 7Networks_LH_Limbic_TempPole_2 | Limbic | Schaefer |
| 305 | 7Networks_LH_Limbic_TempPole_3 | Limbic | Schaefer |
| 306 | 7Networks_LH_Limbic_TempPole_4 | Limbic | Schaefer |
| 307 | 7Networks_LH_Limbic_TempPole_5 | Limbic | Schaefer |
| 308 | 7Networks_LH_Limbic_TempPole_6 | Limbic | Schaefer |
| 309 | 7Networks_LH_Limbic_TempPole_7 | Limbic | Schaefer |
| 310 | 7Networks_LH_Limbic_TempPole_8 | Limbic | Schaefer |
| 311 | 7Networks_LH_Limbic_TempPole_9 | Limbic | Schaefer |

| | | | |
|---|---|---|---|
| 312 | 7Networks_LH_Limbic_TempPole_10 | Limbic | Schaefer |
| 313 | 7Networks_LH_Limbic_TempPole_11 | Limbic | Schaefer |
| 314 | 7Networks_LH_Limbic_TempPole_12 | Limbic | Schaefer |
| 315 | 7Networks_LH_Limbic_TempPole_13 | Limbic | Schaefer |
| 316 | 7Networks_LH_Limbic_TempPole_14 | Limbic | Schaefer |
| 317 | 7Networks_LH_Limbic_TempPole_15 | Limbic | Schaefer |
| 318 | 7Networks_LH_Cont_Par_1 | Cont | Schaefer |
| 319 | 7Networks_LH_Cont_Par_2 | Cont | Schaefer |
| 320 | 7Networks_LH_Cont_Par_3 | Cont | Schaefer |
| 321 | 7Networks_LH_Cont_Par_4 | Cont | Schaefer |
| 322 | 7Networks_LH_Cont_Par_5 | Cont | Schaefer |
| 323 | 7Networks_LH_Cont_Par_6 | Cont | Schaefer |
| 324 | 7Networks_LH_Cont_Par_7 | Cont | Schaefer |
| 325 | 7Networks_LH_Cont_Par_8 | Cont | Schaefer |
| 326 | 7Networks_LH_Cont_Par_9 | Cont | Schaefer |
| 327 | 7Networks_LH_Cont_Par_10 | Cont | Schaefer |
| 328 | 7Networks_LH_Cont_Par_11 | Cont | Schaefer |
| 329 | 7Networks_LH_Cont_Par_12 | Cont | Schaefer |
| 330 | 7Networks_LH_Cont_Par_13 | Cont | Schaefer |
| 331 | 7Networks_LH_Cont_Par_14 | Cont | Schaefer |
| 332 | 7Networks_LH_Cont_Par_15 | Cont | Schaefer |
| 333 | 7Networks_LH_Cont_Temp_1 | Cont | Schaefer |
| 334 | 7Networks_LH_Cont_Temp_2 | Cont | Schaefer |
| 335 | 7Networks_LH_Cont_Temp_3 | Cont | Schaefer |
| 336 | 7Networks_LH_Cont_Temp_4 | Cont | Schaefer |
| 337 | 7Networks_LH_Cont_PFCd_1 | Cont | Schaefer |
| 338 | 7Networks_LH_Cont_PFCl_1 | Cont | Schaefer |
| 339 | 7Networks_LH_Cont_PFCl_2 | Cont | Schaefer |
| 340 | 7Networks_LH_Cont_PFCl_3 | Cont | Schaefer |
| 341 | 7Networks_LH_Cont_PFCl_4 | Cont | Schaefer |
| 342 | 7Networks_LH_Cont_PFCl_5 | Cont | Schaefer |
| 343 | 7Networks_LH_Cont_PFCl_6 | Cont | Schaefer |
| 344 | 7Networks_LH_Cont_PFCl_7 | Cont | Schaefer |
| 345 | 7Networks_LH_Cont_PFCl_8 | Cont | Schaefer |
| 346 | 7Networks_LH_Cont_PFCl_9 | Cont | Schaefer |
| 347 | 7Networks_LH_Cont_PFCl_10 | Cont | Schaefer |
| 348 | 7Networks_LH_Cont_PFCl_11 | Cont | Schaefer |
| 349 | 7Networks_LH_Cont_PFCl_12 | Cont | Schaefer |
| 350 | 7Networks_LH_Cont_PFCl_13 | Cont | Schaefer |
| 351 | 7Networks_LH_Cont_PFCl_14 | Cont | Schaefer |
| 352 | 7Networks_LH_Cont_PFCl_15 | Cont | Schaefer |
| 353 | 7Networks_LH_Cont_PFCl_16 | Cont | Schaefer |
| 354 | 7Networks_LH_Cont_PFCl_17 | Cont | Schaefer |
| 355 | 7Networks_LH_Cont_PFCl_18 | Cont | Schaefer |
| 356 | 7Networks_LH_Cont_PFCl_19 | Cont | Schaefer |

| | | | |
|---|---|---|---|
| 357 | 7Networks_LH_Cont_PFCl_20 | Cont | Schaefer |
| 358 | 7Networks_LH_Cont_OFC_1 | Cont | Schaefer |
| 359 | 7Networks_LH_Cont_PFCv_1 | Cont | Schaefer |
| 360 | 7Networks_LH_Cont_PFCv_2 | Cont | Schaefer |
| 361 | 7Networks_LH_Cont_pCun_1 | Cont | Schaefer |
| 362 | 7Networks_LH_Cont_pCun_2 | Cont | Schaefer |
| 363 | 7Networks_LH_Cont_pCun_3 | Cont | Schaefer |
| 364 | 7Networks_LH_Cont_pCun_4 | Cont | Schaefer |
| 365 | 7Networks_LH_Cont_Cing_1 | Cont | Schaefer |
| 366 | 7Networks_LH_Cont_Cing_2 | Cont | Schaefer |
| 367 | 7Networks_LH_Cont_Cing_3 | Cont | Schaefer |
| 368 | 7Networks_LH_Cont_Cing_4 | Cont | Schaefer |
| 369 | 7Networks_LH_Cont_Cing_5 | Cont | Schaefer |
| 370 | 7Networks_LH_Cont_Cing_6 | Cont | Schaefer |
| 371 | 7Networks_LH_Cont_Cing_7 | Cont | Schaefer |
| 372 | 7Networks_LH_Cont_Cing_8 | Cont | Schaefer |
| 373 | 7Networks_LH_Cont_PFCmp_1 | Cont | Schaefer |
| 374 | 7Networks_LH_Cont_PFCmp_2 | Cont | Schaefer |
| 375 | 7Networks_LH_Default_Par_1 | Default | Schaefer |
| 376 | 7Networks_LH_Default_Par_2 | Default | Schaefer |
| 377 | 7Networks_LH_Default_Par_3 | Default | Schaefer |
| 378 | 7Networks_LH_Default_Par_4 | Default | Schaefer |
| 379 | 7Networks_LH_Default_Par_5 | Default | Schaefer |
| 380 | 7Networks_LH_Default_Par_6 | Default | Schaefer |
| 381 | 7Networks_LH_Default_Par_7 | Default | Schaefer |
| 382 | 7Networks_LH_Default_Par_8 | Default | Schaefer |
| 383 | 7Networks_LH_Default_Par_9 | Default | Schaefer |
| 384 | 7Networks_LH_Default_Par_10 | Default | Schaefer |
| 385 | 7Networks_LH_Default_Par_11 | Default | Schaefer |
| 386 | 7Networks_LH_Default_Par_12 | Default | Schaefer |
| 387 | 7Networks_LH_Default_Par_13 | Default | Schaefer |
| 388 | 7Networks_LH_Default_Par_14 | Default | Schaefer |
| 389 | 7Networks_LH_Default_Par_15 | Default | Schaefer |
| 390 | 7Networks_LH_Default_Par_16 | Default | Schaefer |
| 391 | 7Networks_LH_Default_Par_17 | Default | Schaefer |
| 392 | 7Networks_LH_Default_Par_18 | Default | Schaefer |
| 393 | 7Networks_LH_Default_Par_19 | Default | Schaefer |
| 394 | 7Networks_LH_Default_Temp_1 | Default | Schaefer |
| 395 | 7Networks_LH_Default_Temp_2 | Default | Schaefer |
| 396 | 7Networks_LH_Default_Temp_3 | Default | Schaefer |
| 397 | 7Networks_LH_Default_Temp_4 | Default | Schaefer |
| 398 | 7Networks_LH_Default_Temp_5 | Default | Schaefer |
| 399 | 7Networks_LH_Default_Temp_6 | Default | Schaefer |
| 400 | 7Networks_LH_Default_Temp_7 | Default | Schaefer |
| 401 | 7Networks_LH_Default_Temp_8 | Default | Schaefer |

| | | | |
|---|---|---|---|
| 402 | 7Networks_LH_Default_Temp_9 | Default | Schaefer |
| 403 | 7Networks_LH_Default_Temp_10 | Default | Schaefer |
| 404 | 7Networks_LH_Default_Temp_11 | Default | Schaefer |
| 405 | 7Networks_LH_Default_Temp_12 | Default | Schaefer |
| 406 | 7Networks_LH_Default_Temp_13 | Default | Schaefer |
| 407 | 7Networks_LH_Default_Temp_14 | Default | Schaefer |
| 408 | 7Networks_LH_Default_Temp_15 | Default | Schaefer |
| 409 | 7Networks_LH_Default_Temp_16 | Default | Schaefer |
| 410 | 7Networks_LH_Default_Temp_17 | Default | Schaefer |
| 411 | 7Networks_LH_Default_Temp_18 | Default | Schaefer |
| 412 | 7Networks_LH_Default_Temp_19 | Default | Schaefer |
| 413 | 7Networks_LH_Default_Temp_20 | Default | Schaefer |
| 414 | 7Networks_LH_Default_Temp_21 | Default | Schaefer |
| 415 | 7Networks_LH_Default_Temp_22 | Default | Schaefer |
| 416 | 7Networks_LH_Default_PFC_1 | Default | Schaefer |
| 417 | 7Networks_LH_Default_PFC_2 | Default | Schaefer |
| 418 | 7Networks_LH_Default_PFC_3 | Default | Schaefer |
| 419 | 7Networks_LH_Default_PFC_4 | Default | Schaefer |
| 420 | 7Networks_LH_Default_PFC_5 | Default | Schaefer |
| 421 | 7Networks_LH_Default_PFC_6 | Default | Schaefer |
| 422 | 7Networks_LH_Default_PFC_7 | Default | Schaefer |
| 423 | 7Networks_LH_Default_PFC_8 | Default | Schaefer |
| 424 | 7Networks_LH_Default_PFC_9 | Default | Schaefer |
| 425 | 7Networks_LH_Default_PFC_10 | Default | Schaefer |
| 426 | 7Networks_LH_Default_PFC_11 | Default | Schaefer |
| 427 | 7Networks_LH_Default_PFC_12 | Default | Schaefer |
| 428 | 7Networks_LH_Default_PFC_13 | Default | Schaefer |
| 429 | 7Networks_LH_Default_PFC_14 | Default | Schaefer |
| 430 | 7Networks_LH_Default_PFC_15 | Default | Schaefer |
| 431 | 7Networks_LH_Default_PFC_16 | Default | Schaefer |
| 432 | 7Networks_LH_Default_PFC_17 | Default | Schaefer |
| 433 | 7Networks_LH_Default_PFC_18 | Default | Schaefer |
| 434 | 7Networks_LH_Default_PFC_19 | Default | Schaefer |
| 435 | 7Networks_LH_Default_PFC_20 | Default | Schaefer |
| 436 | 7Networks_LH_Default_PFC_21 | Default | Schaefer |
| 437 | 7Networks_LH_Default_PFC_22 | Default | Schaefer |
| 438 | 7Networks_LH_Default_PFC_23 | Default | Schaefer |
| 439 | 7Networks_LH_Default_PFC_24 | Default | Schaefer |
| 440 | 7Networks_LH_Default_PFC_25 | Default | Schaefer |
| 441 | 7Networks_LH_Default_PFC_26 | Default | Schaefer |
| 442 | 7Networks_LH_Default_PFC_27 | Default | Schaefer |
| 443 | 7Networks_LH_Default_PFC_28 | Default | Schaefer |
| 444 | 7Networks_LH_Default_PFC_29 | Default | Schaefer |
| 445 | 7Networks_LH_Default_PFC_30 | Default | Schaefer |
| 446 | 7Networks_LH_Default_PFC_31 | Default | Schaefer |

| | | | |
|---|---|---|---|
| 447 | 7Networks_LH_Default_PFC_32 | Default | Schaefer |
| 448 | 7Networks_LH_Default_PFC_33 | Default | Schaefer |
| 449 | 7Networks_LH_Default_PFC_34 | Default | Schaefer |
| 450 | 7Networks_LH_Default_PFC_35 | Default | Schaefer |
| 451 | 7Networks_LH_Default_PFC_36 | Default | Schaefer |
| 452 | 7Networks_LH_Default_PFC_37 | Default | Schaefer |
| 453 | 7Networks_LH_Default_PFC_38 | Default | Schaefer |
| 454 | 7Networks_LH_Default_PFC_39 | Default | Schaefer |
| 455 | 7Networks_LH_Default_PFC_40 | Default | Schaefer |
| 456 | 7Networks_LH_Default_PFC_41 | Default | Schaefer |
| 457 | 7Networks_LH_Default_PFC_42 | Default | Schaefer |
| 458 | 7Networks_LH_Default_PFC_43 | Default | Schaefer |
| 459 | 7Networks_LH_Default_PFC_44 | Default | Schaefer |
| 460 | 7Networks_LH_Default_PFC_45 | Default | Schaefer |
| 461 | 7Networks_LH_Default_PFC_46 | Default | Schaefer |
| 462 | 7Networks_LH_Default_PFC_47 | Default | Schaefer |
| 463 | 7Networks_LH_Default_PFC_48 | Default | Schaefer |
| 464 | 7Networks_LH_Default_PFC_49 | Default | Schaefer |
| 465 | 7Networks_LH_Default_PFC_50 | Default | Schaefer |
| 466 | 7Networks_LH_Default_pCunPCC_1 | Default | Schaefer |
| 467 | 7Networks_LH_Default_pCunPCC_2 | Default | Schaefer |
| 468 | 7Networks_LH_Default_pCunPCC_3 | Default | Schaefer |
| 469 | 7Networks_LH_Default_pCunPCC_4 | Default | Schaefer |
| 470 | 7Networks_LH_Default_pCunPCC_5 | Default | Schaefer |
| 471 | 7Networks_LH_Default_pCunPCC_6 | Default | Schaefer |
| 472 | 7Networks_LH_Default_pCunPCC_7 | Default | Schaefer |
| 473 | 7Networks_LH_Default_pCunPCC_8 | Default | Schaefer |
| 474 | 7Networks_LH_Default_pCunPCC_9 | Default | Schaefer |
| 475 | 7Networks_LH_Default_pCunPCC_10 | Default | Schaefer |
| 476 | 7Networks_LH_Default_pCunPCC_11 | Default | Schaefer |
| 477 | 7Networks_LH_Default_pCunPCC_12 | Default | Schaefer |
| 478 | 7Networks_LH_Default_pCunPCC_13 | Default | Schaefer |
| 479 | 7Networks_LH_Default_pCunPCC_14 | Default | Schaefer |
| 480 | 7Networks_LH_Default_pCunPCC_15 | Default | Schaefer |
| 481 | 7Networks_LH_Default_pCunPCC_16 | Default | Schaefer |
| 482 | 7Networks_LH_Default_pCunPCC_17 | Default | Schaefer |
| 483 | 7Networks_LH_Default_pCunPCC_18 | Default | Schaefer |
| 484 | 7Networks_LH_Default_pCunPCC_19 | Default | Schaefer |
| 485 | 7Networks_LH_Default_pCunPCC_20 | Default | Schaefer |
| 486 | 7Networks_LH_Default_pCunPCC_21 | Default | Schaefer |
| 487 | 7Networks_LH_Default_pCunPCC_22 | Default | Schaefer |
| 488 | 7Networks_LH_Default_pCunPCC_23 | Default | Schaefer |
| 489 | 7Networks_LH_Default_pCunPCC_24 | Default | Schaefer |
| 490 | 7Networks_LH_Default_pCunPCC_25 | Default | Schaefer |
| 491 | 7Networks_LH_Default_pCunPCC_26 | Default | Schaefer |

| | | | |
|---|---|---|---|
| 492 | 7Networks_LH_Default_pCunPCC_27 | Default | Schaefer |
| 493 | 7Networks_LH_Default_pCunPCC_28 | Default | Schaefer |
| 494 | 7Networks_LH_Default_pCunPCC_29 | Default | Schaefer |
| 495 | 7Networks_LH_Default_pCunPCC_30 | Default | Schaefer |
| 496 | 7Networks_LH_Default_pCunPCC_31 | Default | Schaefer |
| 497 | 7Networks_LH_Default_pCunPCC_32 | Default | Schaefer |
| 498 | 7Networks_LH_Default_PHC_1 | Default | Schaefer |
| 499 | 7Networks_LH_Default_PHC_2 | Default | Schaefer |
| 500 | 7Networks_LH_Default_PHC_3 | Default | Schaefer |
| 501 | 7Networks_RH_Vis_1 | Vis | Schaefer |
| 502 | 7Networks_RH_Vis_2 | Vis | Schaefer |
| 503 | 7Networks_RH_Vis_3 | Vis | Schaefer |
| 504 | 7Networks_RH_Vis_4 | Vis | Schaefer |
| 505 | 7Networks_RH_Vis_5 | Vis | Schaefer |
| 506 | 7Networks_RH_Vis_6 | Vis | Schaefer |
| 507 | 7Networks_RH_Vis_7 | Vis | Schaefer |
| 508 | 7Networks_RH_Vis_8 | Vis | Schaefer |
| 509 | 7Networks_RH_Vis_9 | Vis | Schaefer |
| 510 | 7Networks_RH_Vis_10 | Vis | Schaefer |
| 511 | 7Networks_RH_Vis_11 | Vis | Schaefer |
| 512 | 7Networks_RH_Vis_12 | Vis | Schaefer |
| 513 | 7Networks_RH_Vis_13 | Vis | Schaefer |
| 514 | 7Networks_RH_Vis_14 | Vis | Schaefer |
| 515 | 7Networks_RH_Vis_15 | Vis | Schaefer |
| 516 | 7Networks_RH_Vis_16 | Vis | Schaefer |
| 517 | 7Networks_RH_Vis_17 | Vis | Schaefer |
| 518 | 7Networks_RH_Vis_18 | Vis | Schaefer |
| 519 | 7Networks_RH_Vis_19 | Vis | Schaefer |
| 520 | 7Networks_RH_Vis_20 | Vis | Schaefer |
| 521 | 7Networks_RH_Vis_21 | Vis | Schaefer |
| 522 | 7Networks_RH_Vis_22 | Vis | Schaefer |
| 523 | 7Networks_RH_Vis_23 | Vis | Schaefer |
| 524 | 7Networks_RH_Vis_24 | Vis | Schaefer |
| 525 | 7Networks_RH_Vis_25 | Vis | Schaefer |
| 526 | 7Networks_RH_Vis_26 | Vis | Schaefer |
| 527 | 7Networks_RH_Vis_27 | Vis | Schaefer |
| 528 | 7Networks_RH_Vis_28 | Vis | Schaefer |
| 529 | 7Networks_RH_Vis_29 | Vis | Schaefer |
| 530 | 7Networks_RH_Vis_30 | Vis | Schaefer |
| 531 | 7Networks_RH_Vis_31 | Vis | Schaefer |
| 532 | 7Networks_RH_Vis_32 | Vis | Schaefer |
| 533 | 7Networks_RH_Vis_33 | Vis | Schaefer |
| 534 | 7Networks_RH_Vis_34 | Vis | Schaefer |
| 535 | 7Networks_RH_Vis_35 | Vis | Schaefer |
| 536 | 7Networks_RH_Vis_36 | Vis | Schaefer |

| | | | |
|---|---|---|---|
| 537 | 7Networks_RH_Vis_37 | Vis | Schaefer |
| 538 | 7Networks_RH_Vis_38 | Vis | Schaefer |
| 539 | 7Networks_RH_Vis_39 | Vis | Schaefer |
| 540 | 7Networks_RH_Vis_40 | Vis | Schaefer |
| 541 | 7Networks_RH_Vis_41 | Vis | Schaefer |
| 542 | 7Networks_RH_Vis_42 | Vis | Schaefer |
| 543 | 7Networks_RH_Vis_43 | Vis | Schaefer |
| 544 | 7Networks_RH_Vis_44 | Vis | Schaefer |
| 545 | 7Networks_RH_Vis_45 | Vis | Schaefer |
| 546 | 7Networks_RH_Vis_46 | Vis | Schaefer |
| 547 | 7Networks_RH_Vis_47 | Vis | Schaefer |
| 548 | 7Networks_RH_Vis_48 | Vis | Schaefer |
| 549 | 7Networks_RH_Vis_49 | Vis | Schaefer |
| 550 | 7Networks_RH_Vis_50 | Vis | Schaefer |
| 551 | 7Networks_RH_Vis_51 | Vis | Schaefer |
| 552 | 7Networks_RH_Vis_52 | Vis | Schaefer |
| 553 | 7Networks_RH_Vis_53 | Vis | Schaefer |
| 554 | 7Networks_RH_Vis_54 | Vis | Schaefer |
| 555 | 7Networks_RH_Vis_55 | Vis | Schaefer |
| 556 | 7Networks_RH_Vis_56 | Vis | Schaefer |
| 557 | 7Networks_RH_Vis_57 | Vis | Schaefer |
| 558 | 7Networks_RH_Vis_58 | Vis | Schaefer |
| 559 | 7Networks_RH_Vis_59 | Vis | Schaefer |
| 560 | 7Networks_RH_Vis_60 | Vis | Schaefer |
| 561 | 7Networks_RH_Vis_61 | Vis | Schaefer |
| 562 | 7Networks_RH_Vis_62 | Vis | Schaefer |
| 563 | 7Networks_RH_Vis_63 | Vis | Schaefer |
| 564 | 7Networks_RH_Vis_64 | Vis | Schaefer |
| 565 | 7Networks_RH_Vis_65 | Vis | Schaefer |
| 566 | 7Networks_RH_Vis_66 | Vis | Schaefer |
| 567 | 7Networks_RH_Vis_67 | Vis | Schaefer |
| 568 | 7Networks_RH_Vis_68 | Vis | Schaefer |
| 569 | 7Networks_RH_Vis_69 | Vis | Schaefer |
| 570 | 7Networks_RH_Vis_70 | Vis | Schaefer |
| 571 | 7Networks_RH_Vis_71 | Vis | Schaefer |
| 572 | 7Networks_RH_Vis_72 | Vis | Schaefer |
| 573 | 7Networks_RH_Vis_73 | Vis | Schaefer |
| 574 | 7Networks_RH_Vis_74 | Vis | Schaefer |
| 575 | 7Networks_RH_Vis_75 | Vis | Schaefer |
| 576 | 7Networks_RH_Vis_76 | Vis | Schaefer |
| 577 | 7Networks_RH_Vis_77 | Vis | Schaefer |
| 578 | 7Networks_RH_Vis_78 | Vis | Schaefer |
| 579 | 7Networks_RH_Vis_79 | Vis | Schaefer |
| 580 | 7Networks_RH_Vis_80 | Vis | Schaefer |
| 581 | 7Networks_RH_Vis_81 | Vis | Schaefer |

| | | | |
|---|---|---|---|
| 582 | 7Networks_RH_SomMot_1 | SomMot | Schaefer |
| 583 | 7Networks_RH_SomMot_2 | SomMot | Schaefer |
| 584 | 7Networks_RH_SomMot_3 | SomMot | Schaefer |
| 585 | 7Networks_RH_SomMot_4 | SomMot | Schaefer |
| 586 | 7Networks_RH_SomMot_5 | SomMot | Schaefer |
| 587 | 7Networks_RH_SomMot_6 | SomMot | Schaefer |
| 588 | 7Networks_RH_SomMot_7 | SomMot | Schaefer |
| 589 | 7Networks_RH_SomMot_8 | SomMot | Schaefer |
| 590 | 7Networks_RH_SomMot_9 | SomMot | Schaefer |
| 591 | 7Networks_RH_SomMot_10 | SomMot | Schaefer |
| 592 | 7Networks_RH_SomMot_11 | SomMot | Schaefer |
| 593 | 7Networks_RH_SomMot_12 | SomMot | Schaefer |
| 594 | 7Networks_RH_SomMot_13 | SomMot | Schaefer |
| 595 | 7Networks_RH_SomMot_14 | SomMot | Schaefer |
| 596 | 7Networks_RH_SomMot_15 | SomMot | Schaefer |
| 597 | 7Networks_RH_SomMot_16 | SomMot | Schaefer |
| 598 | 7Networks_RH_SomMot_17 | SomMot | Schaefer |
| 599 | 7Networks_RH_SomMot_18 | SomMot | Schaefer |
| 600 | 7Networks_RH_SomMot_19 | SomMot | Schaefer |
| 601 | 7Networks_RH_SomMot_20 | SomMot | Schaefer |
| 602 | 7Networks_RH_SomMot_21 | SomMot | Schaefer |
| 603 | 7Networks_RH_SomMot_22 | SomMot | Schaefer |
| 604 | 7Networks_RH_SomMot_23 | SomMot | Schaefer |
| 605 | 7Networks_RH_SomMot_24 | SomMot | Schaefer |
| 606 | 7Networks_RH_SomMot_25 | SomMot | Schaefer |
| 607 | 7Networks_RH_SomMot_26 | SomMot | Schaefer |
| 608 | 7Networks_RH_SomMot_27 | SomMot | Schaefer |
| 609 | 7Networks_RH_SomMot_28 | SomMot | Schaefer |
| 610 | 7Networks_RH_SomMot_29 | SomMot | Schaefer |
| 611 | 7Networks_RH_SomMot_30 | SomMot | Schaefer |
| 612 | 7Networks_RH_SomMot_31 | SomMot | Schaefer |
| 613 | 7Networks_RH_SomMot_32 | SomMot | Schaefer |
| 614 | 7Networks_RH_SomMot_33 | SomMot | Schaefer |
| 615 | 7Networks_RH_SomMot_34 | SomMot | Schaefer |
| 616 | 7Networks_RH_SomMot_35 | SomMot | Schaefer |
| 617 | 7Networks_RH_SomMot_36 | SomMot | Schaefer |
| 618 | 7Networks_RH_SomMot_37 | SomMot | Schaefer |
| 619 | 7Networks_RH_SomMot_38 | SomMot | Schaefer |
| 620 | 7Networks_RH_SomMot_39 | SomMot | Schaefer |
| 621 | 7Networks_RH_SomMot_40 | SomMot | Schaefer |
| 622 | 7Networks_RH_SomMot_41 | SomMot | Schaefer |
| 623 | 7Networks_RH_SomMot_42 | SomMot | Schaefer |
| 624 | 7Networks_RH_SomMot_43 | SomMot | Schaefer |
| 625 | 7Networks_RH_SomMot_44 | SomMot | Schaefer |
| 626 | 7Networks_RH_SomMot_45 | SomMot | Schaefer |

| | | | |
|---|---|---|---|
| 627 | 7Networks_RH_SomMot_46 | SomMot | Schaefer |
| 628 | 7Networks_RH_SomMot_47 | SomMot | Schaefer |
| 629 | 7Networks_RH_SomMot_48 | SomMot | Schaefer |
| 630 | 7Networks_RH_SomMot_49 | SomMot | Schaefer |
| 631 | 7Networks_RH_SomMot_50 | SomMot | Schaefer |
| 632 | 7Networks_RH_SomMot_51 | SomMot | Schaefer |
| 633 | 7Networks_RH_SomMot_52 | SomMot | Schaefer |
| 634 | 7Networks_RH_SomMot_53 | SomMot | Schaefer |
| 635 | 7Networks_RH_SomMot_54 | SomMot | Schaefer |
| 636 | 7Networks_RH_SomMot_55 | SomMot | Schaefer |
| 637 | 7Networks_RH_SomMot_56 | SomMot | Schaefer |
| 638 | 7Networks_RH_SomMot_57 | SomMot | Schaefer |
| 639 | 7Networks_RH_SomMot_58 | SomMot | Schaefer |
| 640 | 7Networks_RH_SomMot_59 | SomMot | Schaefer |
| 641 | 7Networks_RH_SomMot_60 | SomMot | Schaefer |
| 642 | 7Networks_RH_SomMot_61 | SomMot | Schaefer |
| 643 | 7Networks_RH_SomMot_62 | SomMot | Schaefer |
| 644 | 7Networks_RH_SomMot_63 | SomMot | Schaefer |
| 645 | 7Networks_RH_SomMot_64 | SomMot | Schaefer |
| 646 | 7Networks_RH_SomMot_65 | SomMot | Schaefer |
| 647 | 7Networks_RH_SomMot_66 | SomMot | Schaefer |
| 648 | 7Networks_RH_SomMot_67 | SomMot | Schaefer |
| 649 | 7Networks_RH_SomMot_68 | SomMot | Schaefer |
| 650 | 7Networks_RH_SomMot_69 | SomMot | Schaefer |
| 651 | 7Networks_RH_SomMot_70 | SomMot | Schaefer |
| 652 | 7Networks_RH_SomMot_71 | SomMot | Schaefer |
| 653 | 7Networks_RH_SomMot_72 | SomMot | Schaefer |
| 654 | 7Networks_RH_SomMot_73 | SomMot | Schaefer |
| 655 | 7Networks_RH_SomMot_74 | SomMot | Schaefer |
| 656 | 7Networks_RH_SomMot_75 | SomMot | Schaefer |
| 657 | 7Networks_RH_SomMot_76 | SomMot | Schaefer |
| 658 | 7Networks_RH_SomMot_77 | SomMot | Schaefer |
| 659 | 7Networks_RH_SomMot_78 | SomMot | Schaefer |
| 660 | 7Networks_RH_SomMot_79 | SomMot | Schaefer |
| 661 | 7Networks_RH_SomMot_80 | SomMot | Schaefer |
| 662 | 7Networks_RH_SomMot_81 | SomMot | Schaefer |
| 663 | 7Networks_RH_SomMot_82 | SomMot | Schaefer |
| 664 | 7Networks_RH_SomMot_83 | SomMot | Schaefer |
| 665 | 7Networks_RH_SomMot_84 | SomMot | Schaefer |
| 666 | 7Networks_RH_SomMot_85 | SomMot | Schaefer |
| 667 | 7Networks_RH_SomMot_86 | SomMot | Schaefer |
| 668 | 7Networks_RH_SomMot_87 | SomMot | Schaefer |
| 669 | 7Networks_RH_SomMot_88 | SomMot | Schaefer |
| 670 | 7Networks_RH_SomMot_89 | SomMot | Schaefer |
| 671 | 7Networks_RH_SomMot_90 | SomMot | Schaefer |

| | | | |
|---|---|---|---|
| 672 | 7Networks_RH_SomMot_91 | SomMot | Schaefer |
| 673 | 7Networks_RH_SomMot_92 | SomMot | Schaefer |
| 674 | 7Networks_RH_SomMot_93 | SomMot | Schaefer |
| 675 | 7Networks_RH_SomMot_94 | SomMot | Schaefer |
| 676 | 7Networks_RH_SomMot_95 | SomMot | Schaefer |
| 677 | 7Networks_RH_SomMot_96 | SomMot | Schaefer |
| 678 | 7Networks_RH_SomMot_97 | SomMot | Schaefer |
| 679 | 7Networks_RH_SomMot_98 | SomMot | Schaefer |
| 680 | 7Networks_RH_SomMot_99 | SomMot | Schaefer |
| 681 | 7Networks_RH_SomMot_100 | SomMot | Schaefer |
| 682 | 7Networks_RH_SomMot_101 | SomMot | Schaefer |
| 683 | 7Networks_RH_SomMot_102 | SomMot | Schaefer |
| 684 | 7Networks_RH_SomMot_103 | SomMot | Schaefer |
| 685 | 7Networks_RH_DorsAttn_Post_1 | SalVentAt | Schaefer |
| 686 | 7Networks_RH_DorsAttn_Post_2 | SalVentAt | Schaefer |
| 687 | 7Networks_RH_DorsAttn_Post_3 | SalVentAt | Schaefer |
| 688 | 7Networks_RH_DorsAttn_Post_4 | SalVentAt | Schaefer |
| 689 | 7Networks_RH_DorsAttn_Post_5 | SalVentAt | Schaefer |
| 690 | 7Networks_RH_DorsAttn_Post_6 | SalVentAt | Schaefer |
| 691 | 7Networks_RH_DorsAttn_Post_7 | SalVentAt | Schaefer |
| 692 | 7Networks_RH_DorsAttn_Post_8 | SalVentAt | Schaefer |
| 693 | 7Networks_RH_DorsAttn_Post_9 | SalVentAt | Schaefer |
| 694 | 7Networks_RH_DorsAttn_Post_10 | SalVentAt | Schaefer |
| 695 | 7Networks_RH_DorsAttn_Post_11 | SalVentAt | Schaefer |
| 696 | 7Networks_RH_DorsAttn_Post_12 | SalVentAt | Schaefer |
| 697 | 7Networks_RH_DorsAttn_Post_13 | SalVentAt | Schaefer |
| 698 | 7Networks_RH_DorsAttn_Post_14 | SalVentAt | Schaefer |
| 699 | 7Networks_RH_DorsAttn_Post_15 | SalVentAt | Schaefer |
| 700 | 7Networks_RH_DorsAttn_Post_16 | SalVentAt | Schaefer |
| 701 | 7Networks_RH_DorsAttn_Post_17 | SalVentAt | Schaefer |
| 702 | 7Networks_RH_DorsAttn_Post_18 | SalVentAt | Schaefer |
| 703 | 7Networks_RH_DorsAttn_Post_19 | SalVentAt | Schaefer |
| 704 | 7Networks_RH_DorsAttn_Post_20 | SalVentAt | Schaefer |
| 705 | 7Networks_RH_DorsAttn_Post_21 | SalVentAt | Schaefer |
| 706 | 7Networks_RH_DorsAttn_Post_22 | SalVentAt | Schaefer |
| 707 | 7Networks_RH_DorsAttn_Post_23 | SalVentAt | Schaefer |
| 708 | 7Networks_RH_DorsAttn_Post_24 | SalVentAt | Schaefer |
| 709 | 7Networks_RH_DorsAttn_Post_25 | SalVentAt | Schaefer |
| 710 | 7Networks_RH_DorsAttn_Post_26 | SalVentAt | Schaefer |
| 711 | 7Networks_RH_DorsAttn_Post_27 | SalVentAt | Schaefer |
| 712 | 7Networks_RH_DorsAttn_Post_28 | SalVentAt | Schaefer |
| 713 | 7Networks_RH_DorsAttn_Post_29 | SalVentAt | Schaefer |
| 714 | 7Networks_RH_DorsAttn_Post_30 | SalVentAt | Schaefer |
| 715 | 7Networks_RH_DorsAttn_Post_31 | SalVentAt | Schaefer |
| 716 | 7Networks_RH_DorsAttn_Post_32 | SalVentAt | Schaefer |

| | | | |
|---|---|---|---|
| 717 | 7Networks_RH_DorsAttn_Post_33 | SalVentAt | Schaefer |
| 718 | 7Networks_RH_DorsAttn_Post_34 | SalVentAt | Schaefer |
| 719 | 7Networks_RH_DorsAttn_Post_35 | SalVentAt | Schaefer |
| 720 | 7Networks_RH_DorsAttn_Post_36 | SalVentAt | Schaefer |
| 721 | 7Networks_RH_DorsAttn_Post_37 | SalVentAt | Schaefer |
| 722 | 7Networks_RH_DorsAttn_Post_38 | SalVentAt | Schaefer |
| 723 | 7Networks_RH_DorsAttn_Post_39 | SalVentAt | Schaefer |
| 724 | 7Networks_RH_DorsAttn_Post_40 | SalVentAt | Schaefer |
| 725 | 7Networks_RH_DorsAttn_Post_41 | SalVentAt | Schaefer |
| 726 | 7Networks_RH_DorsAttn_Post_42 | SalVentAt | Schaefer |
| 727 | 7Networks_RH_DorsAttn_Post_43 | SalVentAt | Schaefer |
| 728 | 7Networks_RH_DorsAttn_Post_44 | SalVentAt | Schaefer |
| 729 | 7Networks_RH_DorsAttn_Post_45 | SalVentAt | Schaefer |
| 730 | 7Networks_RH_DorsAttn_Post_46 | SalVentAt | Schaefer |
| 731 | 7Networks_RH_DorsAttn_Post_47 | SalVentAt | Schaefer |
| 732 | 7Networks_RH_DorsAttn_Post_48 | SalVentAt | Schaefer |
| 733 | 7Networks_RH_DorsAttn_Post_49 | SalVentAt | Schaefer |
| 734 | 7Networks_RH_DorsAttn_Post_50 | SalVentAt | Schaefer |
| 735 | 7Networks_RH_DorsAttn_Post_51 | SalVentAt | Schaefer |
| 736 | 7Networks_RH_DorsAttn_Post_52 | SalVentAt | Schaefer |
| 737 | 7Networks_RH_DorsAttn_FEF_1 | SalVentAt | Schaefer |
| 738 | 7Networks_RH_DorsAttn_FEF_2 | SalVentAt | Schaefer |
| 739 | 7Networks_RH_DorsAttn_FEF_3 | SalVentAt | Schaefer |
| 740 | 7Networks_RH_DorsAttn_FEF_4 | SalVentAt | Schaefer |
| 741 | 7Networks_RH_DorsAttn_FEF_5 | SalVentAt | Schaefer |
| 742 | 7Networks_RH_DorsAttn_FEF_6 | SalVentAt | Schaefer |
| 743 | 7Networks_RH_DorsAttn_PrCv_1 | SalVentAt | Schaefer |
| 744 | 7Networks_RH_DorsAttn_PrCv_2 | SalVentAt | Schaefer |
| 745 | 7Networks_RH_DorsAttn_PrCv_3 | SalVentAt | Schaefer |
| 746 | 7Networks_RH_SalVentAttn_TempOccPar_1 | DorsAttn | Schaefer |
| 747 | 7Networks_RH_SalVentAttn_TempOccPar_2 | DorsAttn | Schaefer |
| 748 | 7Networks_RH_SalVentAttn_TempOccPar_3 | DorsAttn | Schaefer |
| 749 | 7Networks_RH_SalVentAttn_TempOccPar_4 | DorsAttn | Schaefer |
| 750 | 7Networks_RH_SalVentAttn_TempOccPar_5 | DorsAttn | Schaefer |
| 751 | 7Networks_RH_SalVentAttn_TempOccPar_6 | DorsAttn | Schaefer |
| 752 | 7Networks_RH_SalVentAttn_TempOccPar_7 | DorsAttn | Schaefer |
| 753 | 7Networks_RH_SalVentAttn_TempOccPar_8 | DorsAttn | Schaefer |
| 754 | 7Networks_RH_SalVentAttn_TempOccPar_9 | DorsAttn | Schaefer |
| 755 | 7Networks_RH_SalVentAttn_TempOccPar_10 | DorsAttn | Schaefer |
| 756 | 7Networks_RH_SalVentAttn_TempOccPar_11 | DorsAttn | Schaefer |
| 757 | 7Networks_RH_SalVentAttn_TempOccPar_12 | DorsAttn | Schaefer |
| 758 | 7Networks_RH_SalVentAttn_TempOccPar_13 | DorsAttn | Schaefer |
| 759 | 7Networks_RH_SalVentAttn_TempOccPar_14 | DorsAttn | Schaefer |
| 760 | 7Networks_RH_SalVentAttn_TempOccPar_15 | DorsAttn | Schaefer |
| 761 | 7Networks_RH_SalVentAttn_TempOccPar_16 | DorsAttn | Schaefer |

| | | | |
|---|---|---|---|
| 762 | 7Networks_RH_SalVentAttn_TempOccPar_17 | DorsAttn | Schaefer |
| 763 | 7Networks_RH_SalVentAttn_TempOccPar_18 | DorsAttn | Schaefer |
| 764 | 7Networks_RH_SalVentAttn_PrC_1 | DorsAttn | Schaefer |
| 765 | 7Networks_RH_SalVentAttn_PrC_2 | DorsAttn | Schaefer |
| 766 | 7Networks_RH_SalVentAttn_FrOperIns_1 | DorsAttn | Schaefer |
| 767 | 7Networks_RH_SalVentAttn_FrOperIns_2 | DorsAttn | Schaefer |
| 768 | 7Networks_RH_SalVentAttn_FrOperIns_3 | DorsAttn | Schaefer |
| 769 | 7Networks_RH_SalVentAttn_FrOperIns_4 | DorsAttn | Schaefer |
| 770 | 7Networks_RH_SalVentAttn_FrOperIns_5 | DorsAttn | Schaefer |
| 771 | 7Networks_RH_SalVentAttn_FrOperIns_6 | DorsAttn | Schaefer |
| 772 | 7Networks_RH_SalVentAttn_FrOperIns_7 | DorsAttn | Schaefer |
| 773 | 7Networks_RH_SalVentAttn_FrOperIns_8 | DorsAttn | Schaefer |
| 774 | 7Networks_RH_SalVentAttn_FrOperIns_9 | DorsAttn | Schaefer |
| 775 | 7Networks_RH_SalVentAttn_FrOperIns_10 | DorsAttn | Schaefer |
| 776 | 7Networks_RH_SalVentAttn_FrOperIns_11 | DorsAttn | Schaefer |
| 777 | 7Networks_RH_SalVentAttn_FrOperIns_12 | DorsAttn | Schaefer |
| 778 | 7Networks_RH_SalVentAttn_FrOperIns_13 | DorsAttn | Schaefer |
| 779 | 7Networks_RH_SalVentAttn_FrOperIns_14 | DorsAttn | Schaefer |
| 780 | 7Networks_RH_SalVentAttn_FrOperIns_15 | DorsAttn | Schaefer |
| 781 | 7Networks_RH_SalVentAttn_FrOperIns_16 | DorsAttn | Schaefer |
| 782 | 7Networks_RH_SalVentAttn_FrOperIns_17 | DorsAttn | Schaefer |
| 783 | 7Networks_RH_SalVentAttn_FrOperIns_18 | DorsAttn | Schaefer |
| 784 | 7Networks_RH_SalVentAttn_FrOperIns_19 | DorsAttn | Schaefer |
| 785 | 7Networks_RH_SalVentAttn_FrOperIns_20 | DorsAttn | Schaefer |
| 786 | 7Networks_RH_SalVentAttn_FrOperIns_21 | DorsAttn | Schaefer |
| 787 | 7Networks_RH_SalVentAttn_FrOperIns_22 | DorsAttn | Schaefer |
| 788 | 7Networks_RH_SalVentAttn_FrOperIns_23 | DorsAttn | Schaefer |
| 789 | 7Networks_RH_SalVentAttn_FrOperIns_24 | DorsAttn | Schaefer |
| 790 | 7Networks_RH_SalVentAttn_PFCl_1 | DorsAttn | Schaefer |
| 791 | 7Networks_RH_SalVentAttn_PFCl_2 | DorsAttn | Schaefer |
| 792 | 7Networks_RH_SalVentAttn_Med_1 | DorsAttn | Schaefer |
| 793 | 7Networks_RH_SalVentAttn_Med_2 | DorsAttn | Schaefer |
| 794 | 7Networks_RH_SalVentAttn_Med_3 | DorsAttn | Schaefer |
| 795 | 7Networks_RH_SalVentAttn_Med_4 | DorsAttn | Schaefer |
| 796 | 7Networks_RH_SalVentAttn_Med_5 | DorsAttn | Schaefer |
| 797 | 7Networks_RH_SalVentAttn_Med_6 | DorsAttn | Schaefer |
| 798 | 7Networks_RH_SalVentAttn_Med_7 | DorsAttn | Schaefer |
| 799 | 7Networks_RH_SalVentAttn_Med_8 | DorsAttn | Schaefer |
| 800 | 7Networks_RH_SalVentAttn_Med_9 | DorsAttn | Schaefer |
| 801 | 7Networks_RH_SalVentAttn_Med_10 | DorsAttn | Schaefer |
| 802 | 7Networks_RH_SalVentAttn_Med_11 | DorsAttn | Schaefer |
| 803 | 7Networks_RH_SalVentAttn_Med_12 | DorsAttn | Schaefer |
| 804 | 7Networks_RH_SalVentAttn_Med_13 | DorsAttn | Schaefer |
| 805 | 7Networks_RH_SalVentAttn_Med_14 | DorsAttn | Schaefer |
| 806 | 7Networks_RH_SalVentAttn_Med_15 | DorsAttn | Schaefer |

| | | | |
|---|---|---|---|
| 807 | 7Networks_RH_SalVentAttn_Med_16 | DorsAttn | Schaefer |
| 808 | 7Networks_RH_SalVentAttn_Med_17 | DorsAttn | Schaefer |
| 809 | 7Networks_RH_SalVentAttn_Med_18 | DorsAttn | Schaefer |
| 810 | 7Networks_RH_SalVentAttn_Med_19 | DorsAttn | Schaefer |
| 811 | 7Networks_RH_SalVentAttn_Med_20 | DorsAttn | Schaefer |
| 812 | 7Networks_RH_Limbic_OFC_1 | Limbic | Schaefer |
| 813 | 7Networks_RH_Limbic_OFC_2 | Limbic | Schaefer |
| 814 | 7Networks_RH_Limbic_OFC_3 | Limbic | Schaefer |
| 815 | 7Networks_RH_Limbic_OFC_4 | Limbic | Schaefer |
| 816 | 7Networks_RH_Limbic_OFC_5 | Limbic | Schaefer |
| 817 | 7Networks_RH_Limbic_OFC_6 | Limbic | Schaefer |
| 818 | 7Networks_RH_Limbic_OFC_7 | Limbic | Schaefer |
| 819 | 7Networks_RH_Limbic_OFC_8 | Limbic | Schaefer |
| 820 | 7Networks_RH_Limbic_OFC_9 | Limbic | Schaefer |
| 821 | 7Networks_RH_Limbic_OFC_10 | Limbic | Schaefer |
| 822 | 7Networks_RH_Limbic_OFC_11 | Limbic | Schaefer |
| 823 | 7Networks_RH_Limbic_OFC_12 | Limbic | Schaefer |
| 824 | 7Networks_RH_Limbic_OFC_13 | Limbic | Schaefer |
| 825 | 7Networks_RH_Limbic_OFC_14 | Limbic | Schaefer |
| 826 | 7Networks_RH_Limbic_TempPole_1 | Limbic | Schaefer |
| 827 | 7Networks_RH_Limbic_TempPole_2 | Limbic | Schaefer |
| 828 | 7Networks_RH_Limbic_TempPole_3 | Limbic | Schaefer |
| 829 | 7Networks_RH_Limbic_TempPole_4 | Limbic | Schaefer |
| 830 | 7Networks_RH_Limbic_TempPole_5 | Limbic | Schaefer |
| 831 | 7Networks_RH_Limbic_TempPole_6 | Limbic | Schaefer |
| 832 | 7Networks_RH_Limbic_TempPole_7 | Limbic | Schaefer |
| 833 | 7Networks_RH_Limbic_TempPole_8 | Limbic | Schaefer |
| 834 | 7Networks_RH_Limbic_TempPole_9 | Limbic | Schaefer |
| 835 | 7Networks_RH_Limbic_TempPole_10 | Limbic | Schaefer |
| 836 | 7Networks_RH_Limbic_TempPole_11 | Limbic | Schaefer |
| 837 | 7Networks_RH_Limbic_TempPole_12 | Limbic | Schaefer |
| 838 | 7Networks_RH_Limbic_TempPole_13 | Limbic | Schaefer |
| 839 | 7Networks_RH_Limbic_TempPole_14 | Limbic | Schaefer |
| 840 | 7Networks_RH_Limbic_TempPole_15 | Limbic | Schaefer |
| 841 | 7Networks_RH_Limbic_TempPole_16 | Limbic | Schaefer |
| 842 | 7Networks_RH_Limbic_TempPole_17 | Limbic | Schaefer |
| 843 | 7Networks_RH_Cont_Par_1 | Cont | Schaefer |
| 844 | 7Networks_RH_Cont_Par_2 | Cont | Schaefer |
| 845 | 7Networks_RH_Cont_Par_3 | Cont | Schaefer |
| 846 | 7Networks_RH_Cont_Par_4 | Cont | Schaefer |
| 847 | 7Networks_RH_Cont_Par_5 | Cont | Schaefer |
| 848 | 7Networks_RH_Cont_Par_6 | Cont | Schaefer |
| 849 | 7Networks_RH_Cont_Par_7 | Cont | Schaefer |
| 850 | 7Networks_RH_Cont_Par_8 | Cont | Schaefer |
| 851 | 7Networks_RH_Cont_Par_9 | Cont | Schaefer |

| | | | |
|---|---|---|---|
| 852 | 7Networks_RH_Cont_Par_10 | Cont | Schaefer |
| 853 | 7Networks_RH_Cont_Par_11 | Cont | Schaefer |
| 854 | 7Networks_RH_Cont_Par_12 | Cont | Schaefer |
| 855 | 7Networks_RH_Cont_Par_13 | Cont | Schaefer |
| 856 | 7Networks_RH_Cont_Par_14 | Cont | Schaefer |
| 857 | 7Networks_RH_Cont_Par_15 | Cont | Schaefer |
| 858 | 7Networks_RH_Cont_Par_16 | Cont | Schaefer |
| 859 | 7Networks_RH_Cont_Par_17 | Cont | Schaefer |
| 860 | 7Networks_RH_Cont_Temp_1 | Cont | Schaefer |
| 861 | 7Networks_RH_Cont_Temp_2 | Cont | Schaefer |
| 862 | 7Networks_RH_Cont_Temp_3 | Cont | Schaefer |
| 863 | 7Networks_RH_Cont_Temp_4 | Cont | Schaefer |
| 864 | 7Networks_RH_Cont_PFCv_1 | Cont | Schaefer |
| 865 | 7Networks_RH_Cont_PFCv_2 | Cont | Schaefer |
| 866 | 7Networks_RH_Cont_PFCl_1 | Cont | Schaefer |
| 867 | 7Networks_RH_Cont_PFCl_2 | Cont | Schaefer |
| 868 | 7Networks_RH_Cont_PFCl_3 | Cont | Schaefer |
| 869 | 7Networks_RH_Cont_PFCl_4 | Cont | Schaefer |
| 870 | 7Networks_RH_Cont_PFCl_5 | Cont | Schaefer |
| 871 | 7Networks_RH_Cont_PFCl_6 | Cont | Schaefer |
| 872 | 7Networks_RH_Cont_PFCl_7 | Cont | Schaefer |
| 873 | 7Networks_RH_Cont_PFCl_8 | Cont | Schaefer |
| 874 | 7Networks_RH_Cont_PFCl_9 | Cont | Schaefer |
| 875 | 7Networks_RH_Cont_PFCl_10 | Cont | Schaefer |
| 876 | 7Networks_RH_Cont_PFCl_11 | Cont | Schaefer |
| 877 | 7Networks_RH_Cont_PFCl_12 | Cont | Schaefer |
| 878 | 7Networks_RH_Cont_PFCl_13 | Cont | Schaefer |
| 879 | 7Networks_RH_Cont_PFCl_14 | Cont | Schaefer |
| 880 | 7Networks_RH_Cont_PFCl_15 | Cont | Schaefer |
| 881 | 7Networks_RH_Cont_PFCl_16 | Cont | Schaefer |
| 882 | 7Networks_RH_Cont_PFCl_17 | Cont | Schaefer |
| 883 | 7Networks_RH_Cont_PFCl_18 | Cont | Schaefer |
| 884 | 7Networks_RH_Cont_PFCl_19 | Cont | Schaefer |
| 885 | 7Networks_RH_Cont_PFCl_20 | Cont | Schaefer |
| 886 | 7Networks_RH_Cont_PFCl_21 | Cont | Schaefer |
| 887 | 7Networks_RH_Cont_PFCl_22 | Cont | Schaefer |
| 888 | 7Networks_RH_Cont_PFCl_23 | Cont | Schaefer |
| 889 | 7Networks_RH_Cont_PFCl_24 | Cont | Schaefer |
| 890 | 7Networks_RH_Cont_PFCl_25 | Cont | Schaefer |
| 891 | 7Networks_RH_Cont_PFCl_26 | Cont | Schaefer |
| 892 | 7Networks_RH_Cont_PFCl_27 | Cont | Schaefer |
| 893 | 7Networks_RH_Cont_PFCl_28 | Cont | Schaefer |
| 894 | 7Networks_RH_Cont_PFCl_29 | Cont | Schaefer |
| 895 | 7Networks_RH_Cont_PFCl_30 | Cont | Schaefer |
| 896 | 7Networks_RH_Cont_PFCl_31 | Cont | Schaefer |

| | | | |
|---|---|---|---|
| 897 | 7Networks_RH_Cont_PFCl_32 | Cont | Schaefer |
| 898 | 7Networks_RH_Cont_PFCl_33 | Cont | Schaefer |
| 899 | 7Networks_RH_Cont_PFCl_34 | Cont | Schaefer |
| 900 | 7Networks_RH_Cont_pCun_1 | Cont | Schaefer |
| 901 | 7Networks_RH_Cont_pCun_3 | Cont | Schaefer |
| 902 | 7Networks_RH_Cont_pCun_5 | Cont | Schaefer |
| 903 | 7Networks_RH_Cont_Cing_1 | Cont | Schaefer |
| 904 | 7Networks_RH_Cont_Cing_2 | Cont | Schaefer |
| 905 | 7Networks_RH_Cont_Cing_3 | Cont | Schaefer |
| 906 | 7Networks_RH_Cont_Cing_4 | Cont | Schaefer |
| 907 | 7Networks_RH_Cont_Cing_5 | Cont | Schaefer |
| 908 | 7Networks_RH_Cont_PFCmp_1 | Cont | Schaefer |
| 909 | 7Networks_RH_Cont_PFCmp_2 | Cont | Schaefer |
| 910 | 7Networks_RH_Cont_PFCmp_3 | Cont | Schaefer |
| 911 | 7Networks_RH_Cont_PFCmp_4 | Cont | Schaefer |
| 912 | 7Networks_RH_Cont_PFCmp_5 | Cont | Schaefer |
| 913 | 7Networks_RH_Default_Par_1 | Default | Schaefer |
| 914 | 7Networks_RH_Default_Par_2 | Default | Schaefer |
| 915 | 7Networks_RH_Default_Par_3 | Default | Schaefer |
| 916 | 7Networks_RH_Default_Par_4 | Default | Schaefer |
| 917 | 7Networks_RH_Default_Par_5 | Default | Schaefer |
| 918 | 7Networks_RH_Default_Par_6 | Default | Schaefer |
| 919 | 7Networks_RH_Default_Par_7 | Default | Schaefer |
| 920 | 7Networks_RH_Default_Par_8 | Default | Schaefer |
| 921 | 7Networks_RH_Default_Par_9 | Default | Schaefer |
| 922 | 7Networks_RH_Default_Par_10 | Default | Schaefer |
| 923 | 7Networks_RH_Default_Par_11 | Default | Schaefer |
| 924 | 7Networks_RH_Default_Par_12 | Default | Schaefer |
| 925 | 7Networks_RH_Default_Par_13 | Default | Schaefer |
| 926 | 7Networks_RH_Default_Par_14 | Default | Schaefer |
| 927 | 7Networks_RH_Default_Par_15 | Default | Schaefer |
| 928 | 7Networks_RH_Default_Par_16 | Default | Schaefer |
| 929 | 7Networks_RH_Default_Temp_1 | Default | Schaefer |
| 930 | 7Networks_RH_Default_Temp_2 | Default | Schaefer |
| 931 | 7Networks_RH_Default_Temp_3 | Default | Schaefer |
| 932 | 7Networks_RH_Default_Temp_4 | Default | Schaefer |
| 933 | 7Networks_RH_Default_Temp_5 | Default | Schaefer |
| 934 | 7Networks_RH_Default_Temp_6 | Default | Schaefer |
| 935 | 7Networks_RH_Default_Temp_7 | Default | Schaefer |
| 936 | 7Networks_RH_Default_Temp_8 | Default | Schaefer |
| 937 | 7Networks_RH_Default_Temp_9 | Default | Schaefer |
| 938 | 7Networks_RH_Default_Temp_10 | Default | Schaefer |
| 939 | 7Networks_RH_Default_Temp_11 | Default | Schaefer |
| 940 | 7Networks_RH_Default_Temp_12 | Default | Schaefer |
| 941 | 7Networks_RH_Default_Temp_13 | Default | Schaefer |

| | | | |
|---|---|---|---|
| 942 | 7Networks_RH_Default_Temp_14 | Default | Schaefer |
| 943 | 7Networks_RH_Default_Temp_15 | Default | Schaefer |
| 944 | 7Networks_RH_Default_Temp_16 | Default | Schaefer |
| 945 | 7Networks_RH_Default_Temp_17 | Default | Schaefer |
| 946 | 7Networks_RH_Default_Temp_18 | Default | Schaefer |
| 947 | 7Networks_RH_Default_PFCv_1 | Default | Schaefer |
| 948 | 7Networks_RH_Default_PFCv_2 | Default | Schaefer |
| 949 | 7Networks_RH_Default_PFCv_3 | Default | Schaefer |
| 950 | 7Networks_RH_Default_PFCv_4 | Default | Schaefer |
| 951 | 7Networks_RH_Default_PFCv_5 | Default | Schaefer |
| 952 | 7Networks_RH_Default_PFCv_6 | Default | Schaefer |
| 953 | 7Networks_RH_Default_PFCv_7 | Default | Schaefer |
| 954 | 7Networks_RH_Default_PFCv_8 | Default | Schaefer |
| 955 | 7Networks_RH_Default_PFCv_9 | Default | Schaefer |
| 956 | 7Networks_RH_Default_PFCv_10 | Default | Schaefer |
| 957 | 7Networks_RH_Default_PFCdPFCm_1 | Default | Schaefer |
| 958 | 7Networks_RH_Default_PFCdPFCm_2 | Default | Schaefer |
| 959 | 7Networks_RH_Default_PFCdPFCm_3 | Default | Schaefer |
| 960 | 7Networks_RH_Default_PFCdPFCm_4 | Default | Schaefer |
| 961 | 7Networks_RH_Default_PFCdPFCm_5 | Default | Schaefer |
| 962 | 7Networks_RH_Default_PFCdPFCm_6 | Default | Schaefer |
| 963 | 7Networks_RH_Default_PFCdPFCm_7 | Default | Schaefer |
| 964 | 7Networks_RH_Default_PFCdPFCm_8 | Default | Schaefer |
| 965 | 7Networks_RH_Default_PFCdPFCm_9 | Default | Schaefer |
| 966 | 7Networks_RH_Default_PFCdPFCm_10 | Default | Schaefer |
| 967 | 7Networks_RH_Default_PFCdPFCm_11 | Default | Schaefer |
| 968 | 7Networks_RH_Default_PFCdPFCm_12 | Default | Schaefer |
| 969 | 7Networks_RH_Default_PFCdPFCm_13 | Default | Schaefer |
| 970 | 7Networks_RH_Default_PFCdPFCm_14 | Default | Schaefer |
| 971 | 7Networks_RH_Default_PFCdPFCm_15 | Default | Schaefer |
| 972 | 7Networks_RH_Default_PFCdPFCm_16 | Default | Schaefer |
| 973 | 7Networks_RH_Default_PFCdPFCm_17 | Default | Schaefer |
| 974 | 7Networks_RH_Default_PFCdPFCm_18 | Default | Schaefer |
| 975 | 7Networks_RH_Default_PFCdPFCm_19 | Default | Schaefer |
| 976 | 7Networks_RH_Default_PFCdPFCm_20 | Default | Schaefer |
| 977 | 7Networks_RH_Default_PFCdPFCm_21 | Default | Schaefer |
| 978 | 7Networks_RH_Default_PFCdPFCm_22 | Default | Schaefer |
| 979 | 7Networks_RH_Default_PFCdPFCm_23 | Default | Schaefer |
| 980 | 7Networks_RH_Default_PFCdPFCm_24 | Default | Schaefer |
| 981 | 7Networks_RH_Default_pCunPCC_1 | Default | Schaefer |
| 982 | 7Networks_RH_Default_pCunPCC_2 | Default | Schaefer |
| 983 | 7Networks_RH_Default_pCunPCC_3 | Default | Schaefer |
| 984 | 7Networks_RH_Default_pCunPCC_4 | Default | Schaefer |
| 985 | 7Networks_RH_Default_pCunPCC_5 | Default | Schaefer |
| 986 | 7Networks_RH_Default_pCunPCC_6 | Default | Schaefer |

| | | | |
|---|---|---|---|
| 987 | 7Networks_RH_Default_pCunPCC_7 | Default | Schaefer |
| 988 | 7Networks_RH_Default_pCunPCC_8 | Default | Schaefer |
| 989 | 7Networks_RH_Default_pCunPCC_9 | Default | Schaefer |
| 990 | 7Networks_RH_Default_pCunPCC_10 | Default | Schaefer |
| 991 | 7Networks_RH_Default_pCunPCC_11 | Default | Schaefer |
| 992 | 7Networks_RH_Default_pCunPCC_12 | Default | Schaefer |
| 993 | 7Networks_RH_Default_pCunPCC_13 | Default | Schaefer |
| 994 | 7Networks_RH_Default_pCunPCC_14 | Default | Schaefer |
| 995 | 7Networks_RH_Default_pCunPCC_15 | Default | Schaefer |
| 996 | 7Networks_RH_Default_pCunPCC_16 | Default | Schaefer |
| 997 | 7Networks_RH_Default_pCunPCC_17 | Default | Schaefer |
| 998 | 7Networks_RH_Default_pCunPCC_18 | Default | Schaefer |
| 999 | 7Networks_RH_Cont_pCun_2 | Cont | Schaefer |
| 1000 | 7Networks_RH_Cont_pCun_4 | Cont | Schaefer |
| 1001 | 7Networks_LH_Vis | Vis | Buckner |
| 1002 | 7Networks_LH_SomMot | SomMot | Buckner |
| 1003 | 7Networks_LH_SalVentAt | SalVentAt | Buckner |
| 1004 | 7Networks_LH_DorsAttn | DorsAttn | Buckner |
| 1005 | 7Networks_LH_Limbic | Limbic | Buckner |
| 1006 | 7Networks_LH_Cont | Cont | Buckner |
| 1007 | 7Networks_LH_Default | Default | Buckner |
| 1008 | 7Networks_RH_Vis | Vis | Buckner |
| 1009 | 7Networks_RH_SomMot | SomMot | Buckner |
| 1010 | 7Networks_RH_SalVentAttn | SalVentAt | Buckner |
| 1011 | 7Networks_RH_DorsAttn | DorsAttn | Buckner |
| 1012 | 7Networks_RH_Limbic | Limbic | Buckner |
| 1013 | 7Networks_RH_Cont | Cont | Buckner |
| 1014 | 7Networks_RH_Default | Default | Buckner |
| 1015 | 7Networks_LH_SomMot | SomMot | Choi |
| 1016 | 7Networks_LH_DorsAttn | DorsAttn | Choi |
| 1017 | 7Networks_LH_Limbic | Limbic | Choi |
| 1018 | 7Networks_LH_Cont | Cont | Choi |
| 1019 | 7Networks_LH_Default | Default | Choi |
| 1020 | 7Networks_RH_SomMot | SomMot | Choi |
| 1021 | 7Networks_RH_SalVentAttn | SalVentAt | Choi |
| 1022 | 7Networks_RH_DorsAttn | DorsAttn | Choi |
| 1023 | 7Networks_RH_Limbic | Limbic | Choi |
| 1024 | 7Networks_RH_Cont | Cont | Choi |
| 1025 | 7Networks_RH_Default | Default | Choi |
| 1026 | 7Networks_LH_Vis | Vis | Thalamic |
| 1027 | 7Networks_LH_SomMot | SomMot | Thalamic |
| 1028 | 7Networks_LH_SalVentAt | SalVentAt | Thalamic |
| 1029 | 7Networks_LH_DorsAttn | DorsAttn | Thalamic |
| 1030 | 7Networks_LH_Limbic | Limbic | Thalamic |
| 1031 | 7Networks_LH_Cont | Cont | Thalamic |

| | | | |
|---|---|---|---|
| 1032 | 7Networks_LH_Default | Default | Thalamic |
| 1033 | 7Networks_RH_Vis | Vis | Thalamic |
| 1034 | 7Networks_RH_SomMot | SomMot | Thalamic |
| 1035 | 7Networks_RH_SalVentAttn | SalVentAt | Thalamic |
| 1036 | 7Networks_RH_DorsAttn | DorsAttn | Thalamic |
| 1037 | 7Networks_RH_Limbic | Limbic | Thalamic |
| 1038 | 7Networks_RH_Cont | Cont | Thalamic |
| 1039 | 7Networks_RH_Default | Default | Thalamic |

**Figure S1**

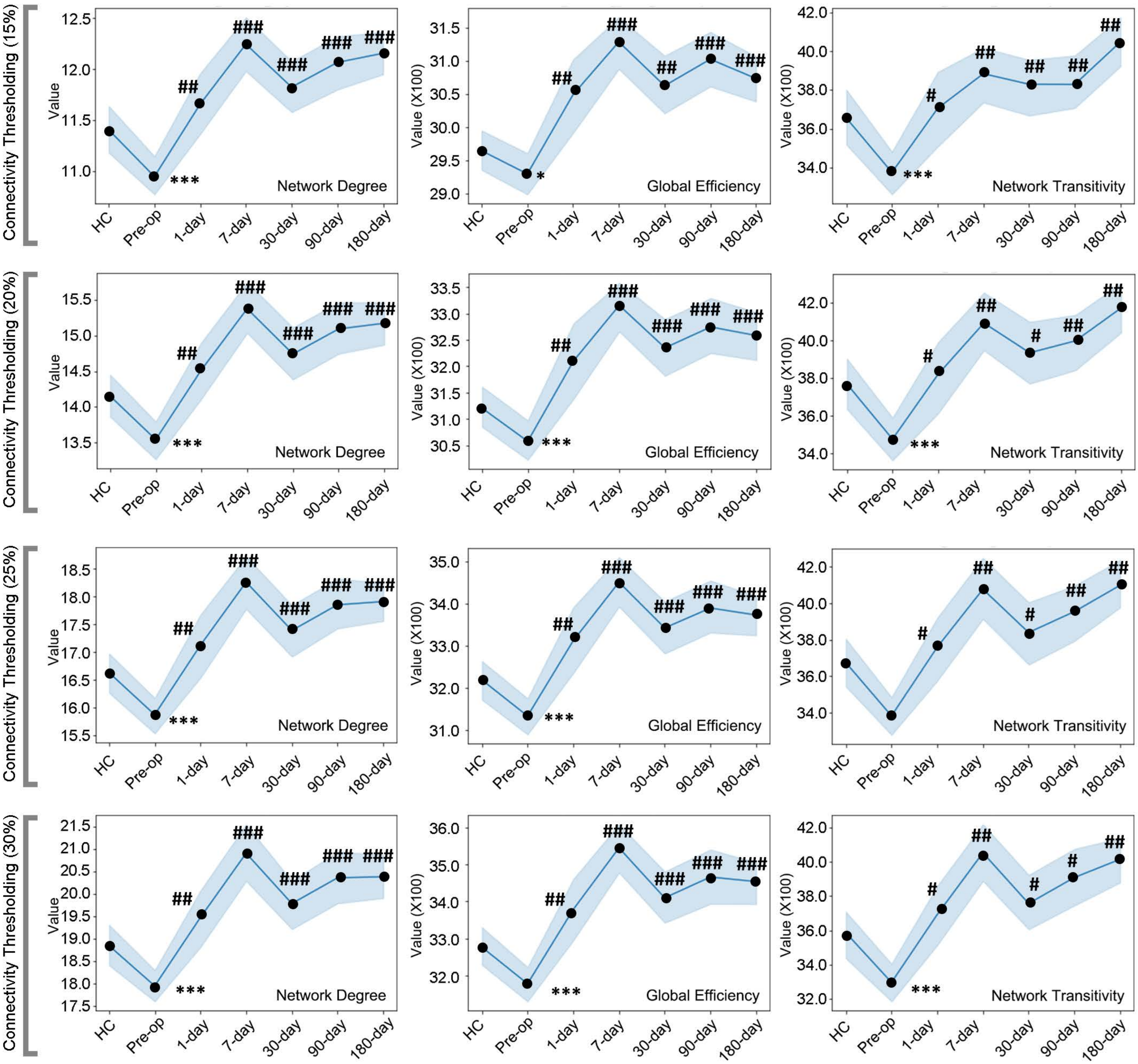


**Figure S1 Temporal changes of hemisphere lateralization for MRgFUS thalamotomy subnetwork under different connectivity matrix thresholding (15-30%).**

(Pre-op = pre-operative; 1-day = 1-day postoperatively; 7-day = 7-day postoperatively; 30-day = 30-day postoperatively; 90-day = 90-day postoperatively; 180-day = 180-day postoperatively. Comparison with match HCs: $^{*}P < 0.05$, $^{***}P < 0.001$; comparison with Pre-op: $^{\#}P < 0.05$, $^{\#\#}P < 0.01$, $^{\#\#}P < 0.001$.).
